# A Standard Framework for Converting Coronary Angiography Reports into Machine-Readable Format Using Large Language Models

**DOI:** 10.1101/2025.05.03.25326846

**Authors:** Ji Woo Song, Ji Yong Jang, Hyeongsoo Kim, Young-Guk Ko, Seng Chan You

## Abstract

**Background and Objectives:** Coronary angiography (CAG) reports contain many details about coronary anatomy, lesion characteristics, and interventional procedures. However, their free-text format limits their research utility. Therefore, we sought to develop and validate a framework leveraging large language models (LLMs) to convert CAG reports automatically into a standardized structured format.

**Methods:** Using 50 CAG reports from a tertiary hospital, we developed a multi-step framework to standardize and extract key information from CAG reports. First, a standard annotation schema was developed by cardiologists. Thereafter, an LLM (GPT-4o) converted the free-text CAG reports into the hierarchical annotation schema in a standardized format. Finally, clinically relevant information was extracted from the standardized schema. One hundred CAG reports from each of two hospitals were used for internal and external test, respectively. The 12 key information points included four CAG-related (previous stent information, lesion characteristics, and anatomical diagnosis) and eight percutaneous coronary intervention (PCI)-related key points (complex PCI criteria and current stent information). For internal test, two interventional cardiologists independently extracted information, with discrepancies resolved through consensus, as reference standard.

**Results:** Based on the reference standard, the proposed framework demonstrated superior accuracy for CAG-related (99.5% vs. 91.8%; p < 0.001) and comparable accuracy for PCI-related key points (98.3% vs. 97.4%; p = 0.512) in the internal test. External test confirmed high accuracy for both CAG-(96.2%) and PCI-related key points (99.4%).

**Conclusions:** This framework demonstrated excellent accuracy in standardizing free-text CAG reports, potentially enabling more efficient utilization of detailed clinical data for cardiovascular research.

**Author’s Summary:** The novel framework that standardizes CAG report is a practical solution to a significant challenge in cardiovascular research — the efficient utilization of detailed procedural and anatomical information untapped in free-text CAG reports. Our framework could enable systematic analysis of large-scale coronary intervention outcomes, reduce the burden of cardologists’ clinical trial recruitment, and support evidence-based clinical decision-making.

## Introduction

Coronary angiography (CAG) reports contain information about coronary artery disease and percutaneous coronary intervention (PCI), providing invaluable data for clinical research. With continued growth in the secondary use of clinical data, observational studies on coronary artery diseases using electronic health records (EHR) continue to increase. However, many of these studies are conducted without the use of detailed information on complex coronary anatomy or procedures.^1–3^ CAG reports are limited by their unstructured, free-text format, which makes it difficult to search, analyze, and process data consistently.^4^

To standardize complex medical information in free-text medical records, we have previously proposed Staged Optimization of Curation, Regularization, and Annotation of clinical text (SOCRATex). This framework initially requires domain experts to define a standardized schema that specifies how clinical information should be organized. Thereafter, experts manually review clinical notes and annotate relevant information according to this predefined schema to create standardized, machine-readable data.^5^ Although this systematic approach effectively converts unstructured clinical notes into analyzable data, it involves a time-consuming manual annotation process and requires significant expert involvement, hampering its large-scale implementation.^6^

Recent advancements in large language models (LLMs), such as ChatGPT, a type of artificial intelligence (AI) used for understanding and processing free-text, can structure vast amounts of free-text within EHRs with minimal programming effort.^7^ Various studies have reported using LLMs to convert free-text radiology and pathology reports into structured formats, highly accurately.^8–10^ However, LLM application in transforming cardiology-related procedure reports into machine-readable formats, particularly CAG reports with their complex procedural details, remains unexplored.

Therefore, building upon our previous work on hierarchical annotation, this study sought to develop a standard framework for the automated transformation of CAG reports into machine-readable formats using an LLM, based on hierarchical annotation. We then compared the accuracy of the extracted information from this machine-readable format with the accuracy of manual review by cardiologists.

## METHODS

### DATASET

CAG studies of adults (>18-years-old), performed from January 1, 2009, to December 31, 2023 (15 years), at Severance Hospital, a tertiary hospital, were retrieved. From these, 50 and 100 CAG reports were randomly selected for the training and internal test sets, respectively. Following the Society for Cardiovascular Angiography and Interventions’ expert consensus,^11^ these CAG reports included the coronary anatomy and described lesions with their location, severity, and morphological characteristics. PCI-related information, including the types and sizes of catheters, balloons, stents, and adjuvant devices, was thoroughly detailed in the CAG report. All reports were written in English, with rare exceptions of Korean texts describing emergency situations during procedures.

Furthermore, 100 random CAG reports from the National Health Insurance Service Ilsan Hospital, a secondary hospital, obtained between January 1, 2023, and December 31, 2023, were retrieved as an external test set.

This study was approved by the Institutional Review Board (IRB) of Severance Hospital (IRB No. 2024-1928-002), Seoul, Korea, and the IRB of the National Health Insurance Service Ilsan Hospital (IRB No. 2024-09-006). The requirement for informed patient consent was waived by the IRBs due to the retrospective nature of the study.

### FRAMEWORK OVERVIEW

Figure 1 illustrates our CAG report standardization framework. After development of a hierarchical annotation schema by experience cardiologists (detailed below), the framework consisted of standardization and extraction steps. In the standardization step, an LLM converted free-text CAG reports into a standardized format based on the expert-defined schema. Thereafter, an automated extraction algorithm extracted the required clinical information. Figure 2 details the development and validation of the framework, including iterative refinement with interventional cardiologists to ensure clinical accuracy and reliability.

**Figure 1.**
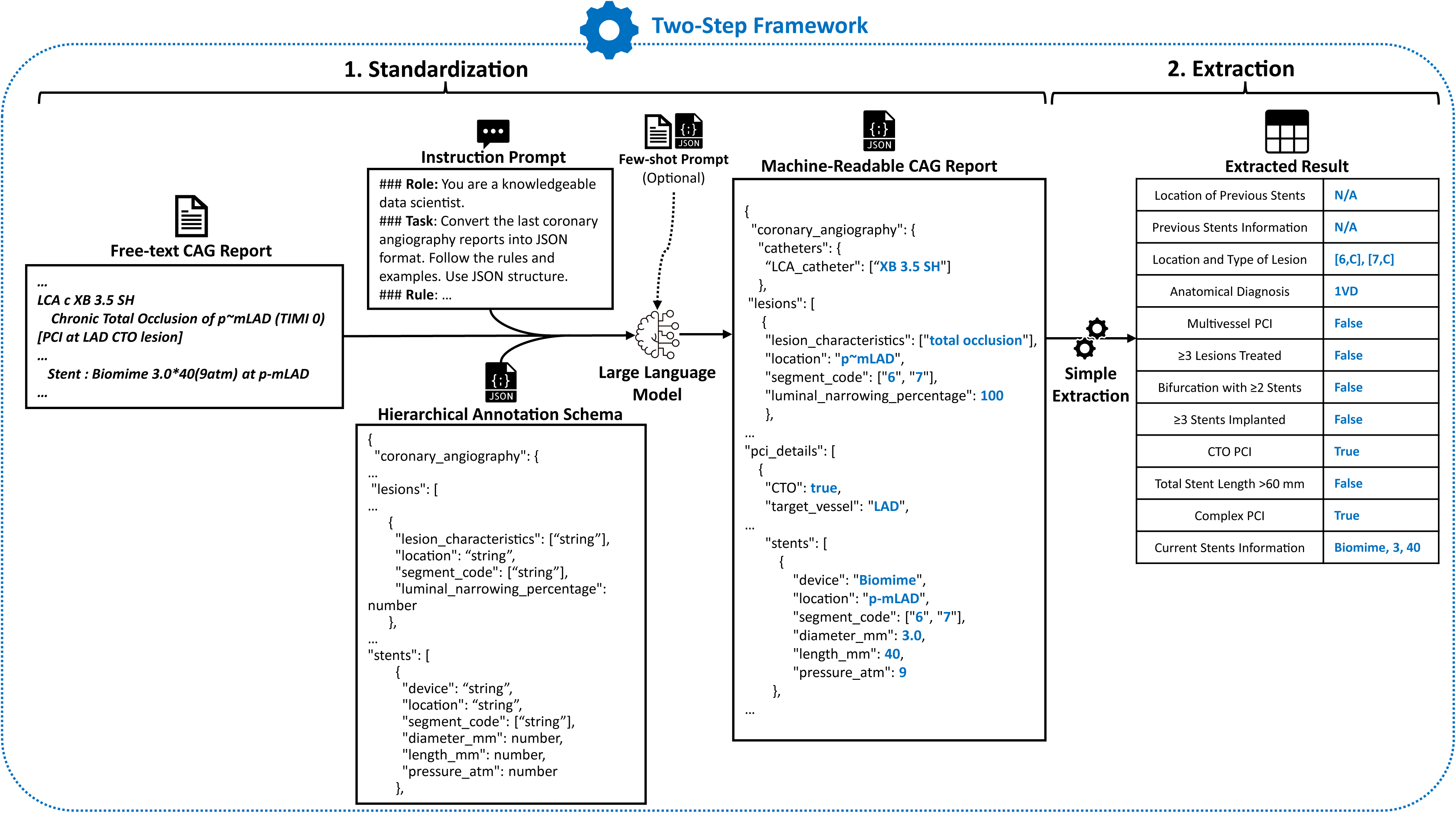
Overview of the two-step framework for free-text CAG report standardization and data extraction. The framework consists of two primary steps: (1) standardization and (2) extraction. In the standardization step, free-text coronary angiography (CAG) reports are converted into a machine-readable hierarchical annotation schema using large language models (LLMs), guided by instruction prompts and optional few-shot prompts. This schema defines the structure and content of the machine-readable format generated in this manner. In the extraction step, 12 key data points are extracted from these machine-readable reports to enable automated and precise data analysis, providing insights into clinical variables, such as lesion characteristics, stent details, and complex percutaneous coronary intervention information.

**Figure 2.**
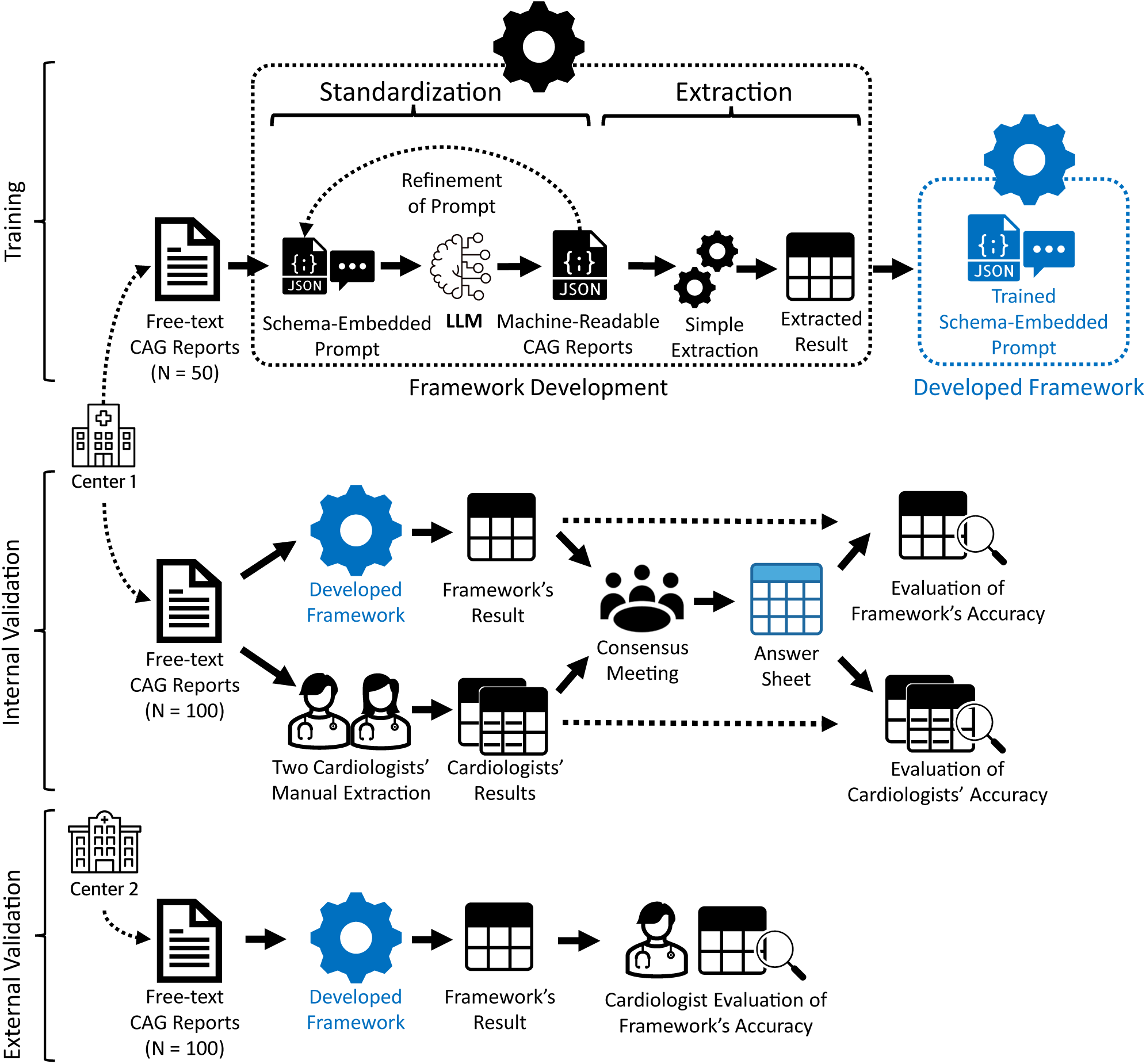
Overview of the training, internal test, and external test processes of the framework. Fifty and 100 reports from electronic health records of Center 1 were used for training the framework and for evaluating the accuracy of the framework and that of the manual extraction of two cardiologists in an internal validation test, respectively. One hundred reports from electronic health records of Center 2 were used as an external test dataset to evaluate the generalizability of the framework. Center 1, Severance Hospital; Center 2, Ilsan Hospital. LLM, large language model

In accordance with the Minimum Reporting Items for Clear Evaluation of Accuracy Reports of Large Language Models in Healthcare (MINI-CLEAR-LLM) checklist, we disclose the details of this study to ensure transparency in utilizing LLMs for healthcare applications in Supplementary Method 1.^12^

### DEVELOPMENT OF THE HIERARCHICAL ANNOTATION SCHEMA

A domain-specific hierarchical annotation schema was collaboratively developed by interventional cardiologists (JYJ) and clinical informatics experts (JWS and SCY). This schema defined how to organize key clinical information from CAG reports systematically, including coronary anatomy, lesion characteristics, and procedural details. Through iterative testing using the training dataset, we refined the schema to ensure comprehensive and accurate capture of complex clinical information. The schema was implemented in JavaScript Object Notation (JSON) format, which enabled efficient organization of multiple clinical events (e.g., multiple lesions or procedures) and flexible representation of clinical details at various levels of granularity.^8–10^ The full annotation schema is shown in Supplementary Figure 1.

### DOCUMENT-LEVEL HIERARCHICAL ANNOTATION

We developed a prompt^13^ to encode domain-specific knowledge, including a Synergy Between Percutaneous Coronary Intervention with Taxus and Cardiac Surgery (SYNTAX) score segmentation system^14^ for precise documentation of coronary anatomy; lesion morphology and characteristics based on American College of Cardiology/American Heart Association (ACC/AHA) classification^15–17^; and details of the intervention, procedural complications, and outcomes.

We utilized Generative Pretrained Transformer 4 Omni (GPT-4o) as the LLM.^18^ The free-text conversion process was initially validated using representative cases from the training dataset that encompassed various clinical scenarios, from simple single-vessel disease to complex multivessel interventions. Among them, eight particularly instructive cases were selected to serve as reference examples, ensuring consistency in the interpretation of complex anatomical and procedural details. While these reference cases were utilized for an internal test, they were not applied in the external test, to avoid institutional bias in reporting styles. The full instruction prompt and few-shot prompts are shown in Supplementary Figure 1 and Supplementary Figure 2, respectively.

### EXTRACTION AND VALIDATION

The extraction process was designed to capture clinically relevant information from the standardized text, with a particular focus on two main categories: CAG-related and PCI-related key information points.

For validation test, we pre-defined the following 12 key information points in the extraction process: four CAG-related key points and eight PCI-related key points. The four CAG-related key points included the location of previous stents, previous stent information, the location and type of lesion, and anatomical diagnosis (e.g., two-vessel disease). *Previous stent information* included the device name, diameter, and length of previous stents. *Type of lesion* (A, B1, B2, and C) was determined for each lesion according to the ACC/AHA classification.

The eight PCI-related key points focused on the six criteria of complex PCI^19^: Multivessel PCI, implantation of ≥ 3 stents; treatment of ≥ 3 lesions; bifurcation PCI using ≥ 2 stents; total stent length > 60 mm; chronic total occlusion (CTO) as the target lesion^20–24^; complex PCI; and current stent information. *Complex PCI* was defined when any one or more criteria were met in the index PCI. *Current stent information* included the device name, diameter, and length of current stents.

The framework’s accuracy was assessed through both internal and external test. For internal test, two experienced cardiologists (JYJ and HSK) independently extracted the 12 key information points manually. Any discrepancies between the framework’s output and manual review were resolved through discussion to consensus, to establish a reference standard. The framework’s performance was compared with that of the cardiologists using Fisher’s exact test (p-value threshold of 0.05). For external test, cardiologists directly evaluated the framework’s extraction results through thorough inspection. The complete extraction algorithm is explained in Supplementary Method 2 and is available at our public repository: https://github.com/jiuisdisciple/CAGtoJSON.

## RESULTS

### COMPOSITION OF TRAINING AND TEST DATASETS

Table 1 summarizes the number of reports included in the training, internal test, and external test datasets, based on the year of report writing and whether they included CAG and PCI procedures. In each CAG report, CAG-related and PCI-related information were intermingled. Therefore, the training set of 50 reports included 47 CAG and 27 PCI cases. Among the 100 reports included in the internal test set were 96 CAG and 44 PCI cases. The 100 reports in the external test dataset included 98 CAG and 58 PCI cases.

**Table 1.** Characteristics of the training, internal test, and external test datasets.

| **Characteristics of the Training, Internal Test and External Test Set** |  |  |  |
| --- | --- | --- | --- |
|  | Training Set | Internal Test Set | External Test Set |
| Total Number of Reports | 50 | 100 | 100 |
| Year of Procedure |  |  |  |
| 2009-2013 | 10 | 25 | 0 |
| 2014-2018 | 22 | 44 | 0 |
| 2019-2023 | 18 | 31 | 0 |
| 2024 | 0 | 0 | 100 |
| Included Procedure |  |  |  |
| CAG and PCI | 26 | 41 | 57 |
| CAG only | 21 | 55 | 41 |
| PCI only | 1 | 3 | 1 |
| Others | 2* | 1* | 1† |

CAG = coronary angiography; PCI = percutaneous coronary intervention.
\* Reports about angiography of coronary artery bypass graft; † Report about inferior vena cava filter placement.

### INTERNAL TEST

Table 2 and Figure 3 show the accuracy of the framework and the mean accuracy of the two cardiologists in extracting clinical key points from CAG reports in the internal test dataset.

**Figure 3.**
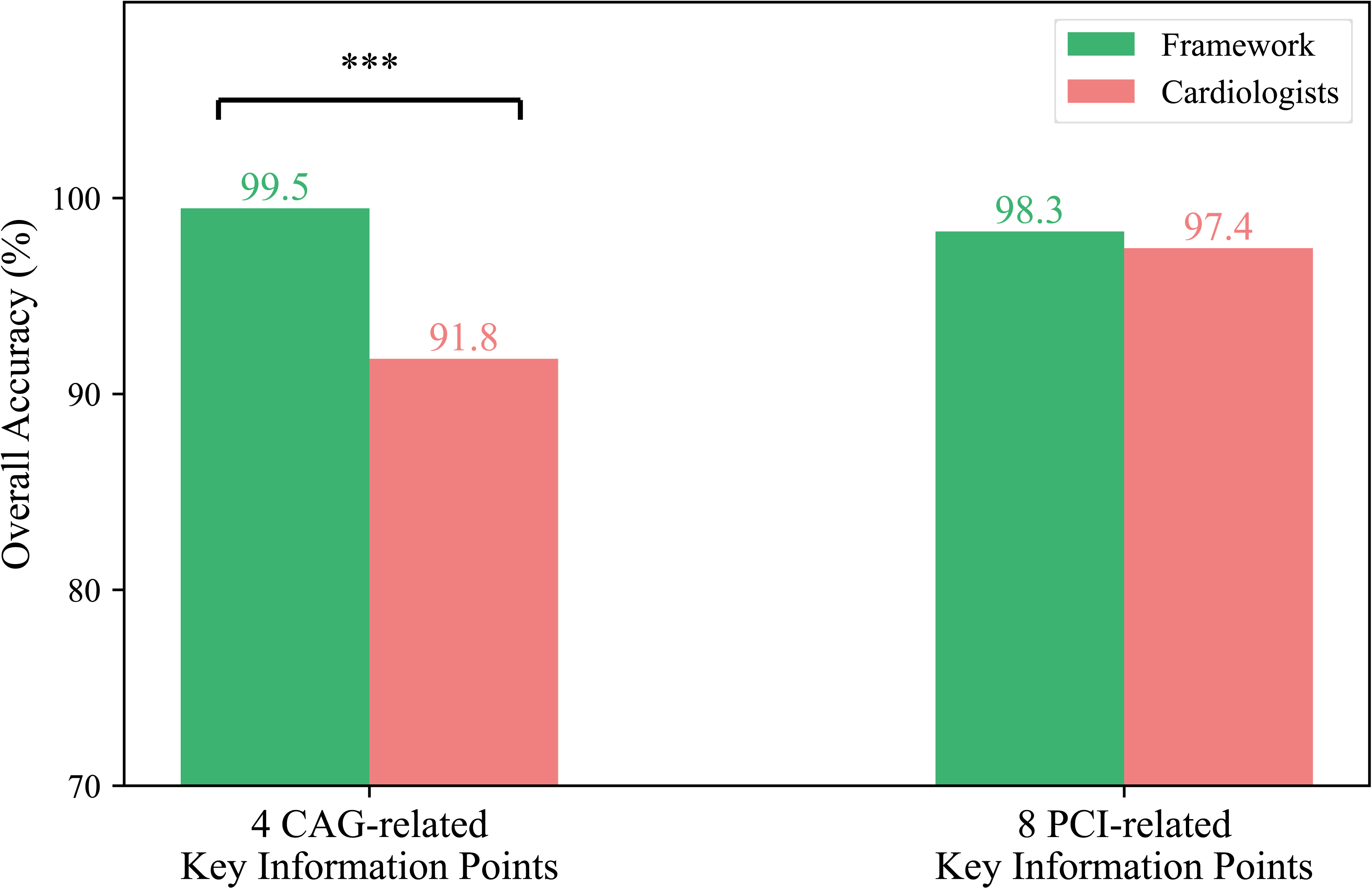
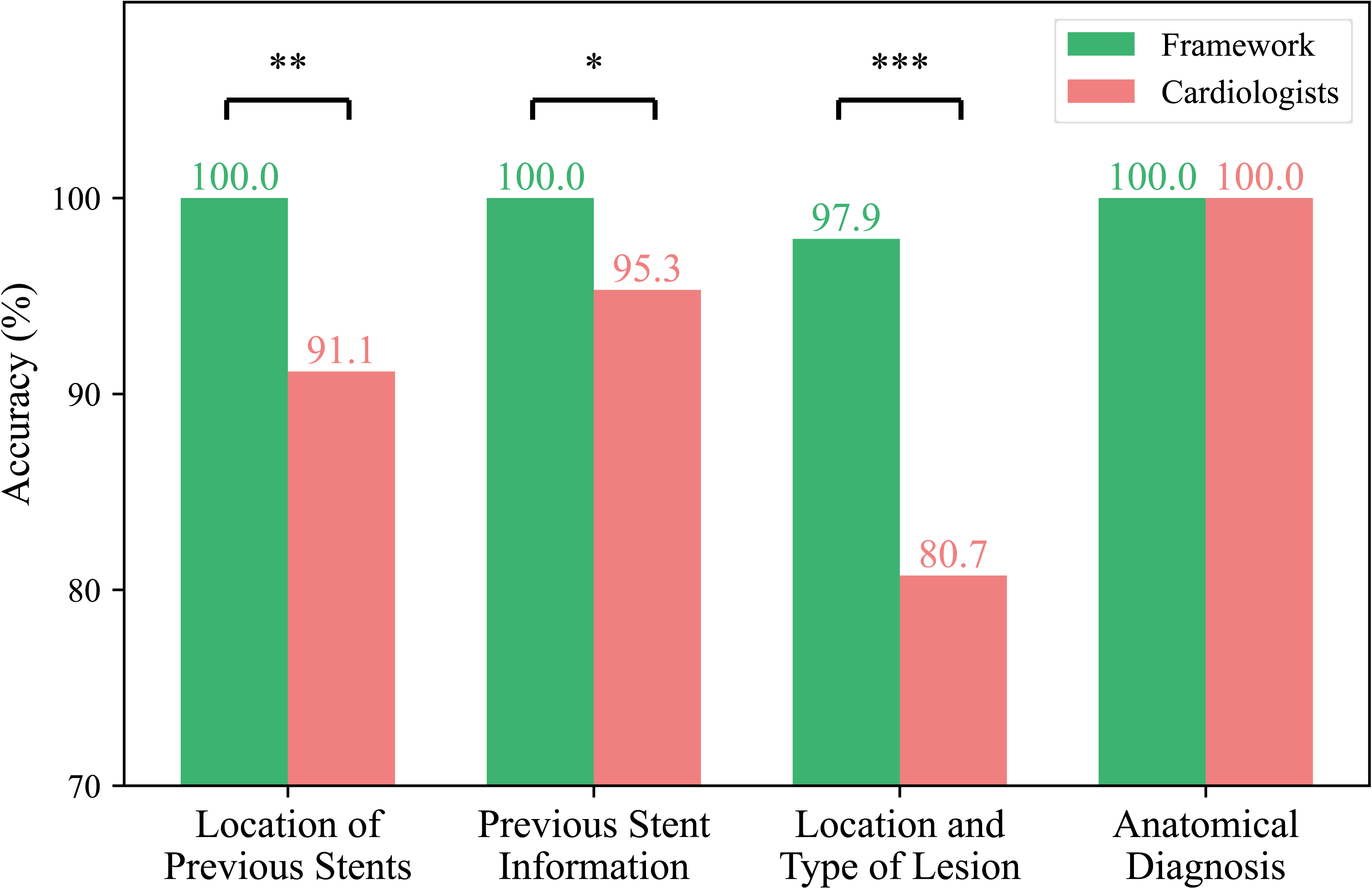
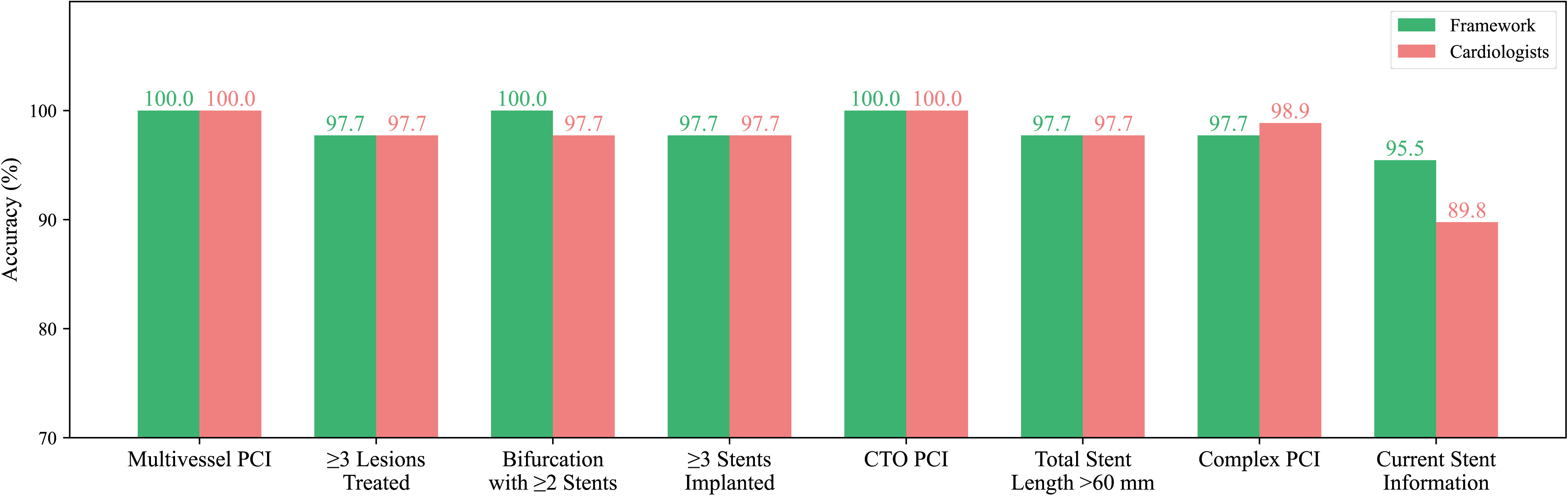
Bar plot showing the accuracy percentages with 95% confidence intervals for the analysis of 12 key information points by the framework and by two cardiologists in the internal validation process. This figure compares the accuracy of the framework and of the two cardiologists in analyzing 12 key information points during the internal validation process. (A) shows the overall accuracy for four coronary angiography (CAG)-related and eight percutaneous coronary intervention (PCI)-related key points. (B) presents individual accuracy for the four CAG-related key points. (C) displays the accuracy for the eight individual PCI-related key points. Statistical significance is indicated as follows: * for p < 0.05, ** for p < 0.01, and *** for p < 0.001.

**Table 2.**
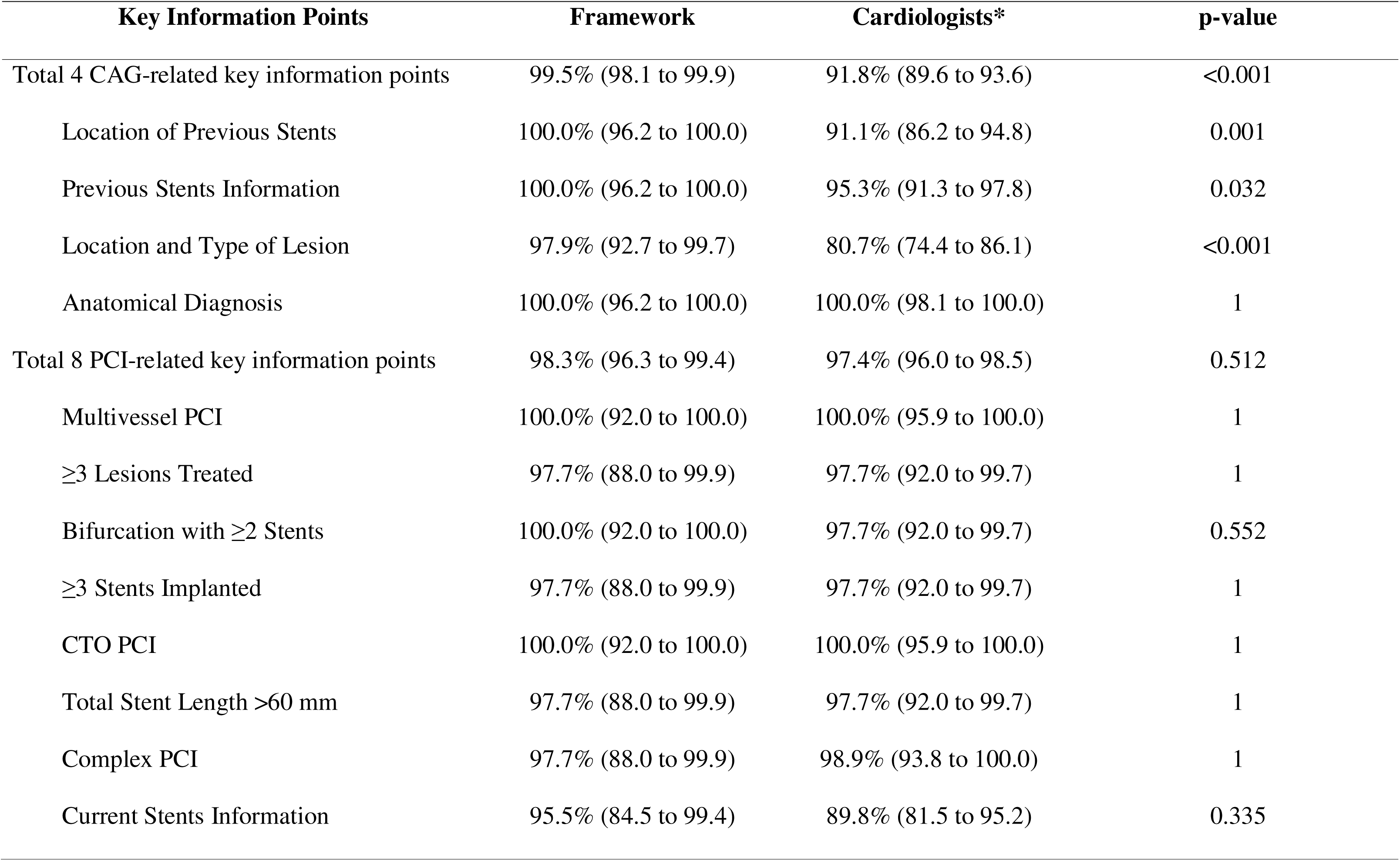
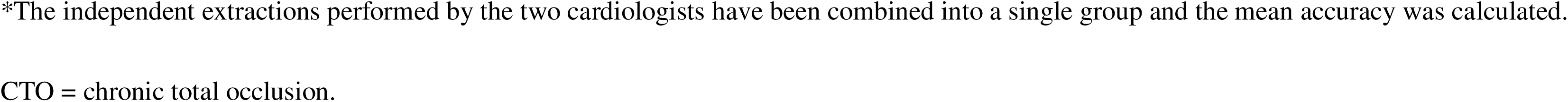
Accuracy percentages with 95% confidence intervals for the analysis of 12 key information points by the framework and by two cardiologists in the internal test.

| Key Information Points | Framework | Cardiologists* | p-value |
| --- | --- | --- | --- |
| Total 4 CAG-related key information points | 99.5% (98.1 to 99.9) | 91.8% (89.6 to 93.6) | <0.001 |
| Location of Previous Stents | 100.0% (96.2 to 100.0) | 91.1% (86.2 to 94.8) | 0.001 |
| Previous Stents Information | 100.0% (96.2 to 100.0) | 95.3% (91.3 to 97.8) | 0.032 |
| Location and Type of Lesion | 97.9% (92.7 to 99.7) | 80.7% (74.4 to 86.1) | <0.001 |
| Anatomical Diagnosis | 100.0% (96.2 to 100.0) | 100.0% (98.1 to 100.0) | 1 |
| Total 8 PCI-related key information points | 98.3% (96.3 to 99.4) | 97.4% (96.0 to 98.5) | 0.512 |
| Multivessel PCI | 100.0% (92.0 to 100.0) | 100.0% (95.9 to 100.0) | 1 |
| ≥3 Lesions Treated | 97.7% (88.0 to 99.9) | 97.7% (92.0 to 99.7) | 1 |
| Bifurcation with ≥2 Stents | 100.0% (92.0 to 100.0) | 97.7% (92.0 to 99.7) | 0.552 |
| ≥3 Stents Implanted | 97.7% (88.0 to 99.9) | 97.7% (92.0 to 99.7) | 1 |
| CTO PCI | 100.0% (92.0 to 100.0) | 100.0% (95.9 to 100.0) | 1 |
| Total Stent Length >60 mm | 97.7% (88.0 to 99.9) | 97.7% (92.0 to 99.7) | 1 |
| Complex PCI | 97.7% (88.0 to 99.9) | 98.9% (93.8 to 100.0) | 1 |
| Current Stents Information | 95.5% (84.5 to 99.4) | 89.8% (81.5 to 95.2) | 0.335 |

\*The independent extractions performed by the two cardiologists have been combined into a single group and the mean accuracy was calculated.
CTO = chronic total occlusion.

The framework demonstrated a significantly higher mean accuracy than that of the cardiologists in extracting the four CAG-related key points (99.5% vs. 91.8%, p < 0.001). For individual CAG key points, the framework showed superior accuracy in *location of previous stents* (100.0% vs. 91.1%, p = 0.001), *previous stent information* (100.0% vs. 95.3%, p = 0.032), and *location and type of lesion* (97.9% vs. 80.7%, p < 0.001). Both the framework and cardiologists achieved 100% accuracy for *anatomical diagnosis*.

For the eight PCI-related key points, the framework showed comparable accuracy to the cardiologists across all items, with mean accuracy scores of 98.3% vs. 97.4% (p = 0.512). The framework and the cardiologists performed similarly for each specific point: *multivessel PCI* (100.0% vs. 100.0%), ≥ *3 lesions treated* (97.7% vs. 97.7%), *bifurcation PCI with* ≥ *2 stents* (100.0% vs. 97.7%), ≥ *3 stents implanted* (97.7% vs. 97.7%), *CTO PCI* (100.0% vs. 100.0%), *total stent length > 60 mm* (97.7% vs. 97.7%), *complex PCI* (97.7% vs. 98.9%), and *current stent information* (95.5% vs. 89.8%). All p-values were above 0.05, indicating no significant differences between the framework and the cardiologists. More detailed results are shown in Supplementary Table 1.

We performed a qualitative analysis to understand the significant discrepancies in accuracy between the framework and the cardiologists. As shown in Supplementary Table 2, cardiologists often generated errors by mislabeling the location of previous stents, current stents, and lesions. They also frequently misclassified the type of lesions due to missed key descriptors, such as “eccentric” and “ostium”. In contrast, the framework consistently performed precise extraction of information from very complex cases, as shown in Supplementary Figure 3A and 3B

### EXTERNAL TEST

Table 3 shows the accuracy of the framework in extracting clinical key points from CAG reports in the external test dataset. For the four CAG-related key points, the framework achieved a mean accuracy of 96.2%, showing consistently high performance across each item. For the eight PCI-related key points, the framework achieved a mean accuracy of 99.4%, also showing consistently high performance across each item. These results indicated the framework’s robust accuracy in extracting key details from both CAG and PCI data. Supplementary Table 3 shows more detailed results.

**Table 3.** Accuracy percentages for the analysis of framework in the external validation test. 12 key information points by the framework in the external validation test.

| **Key Information Points** | **Accuracy of Framework** |
| --- | --- |
| Total 4 CAG-related key information points | 96.2% |
| Location of Previous Stents | 95.9% |
| Previous Stents Information | 98.0% |
| Location and Type of Lesion | 94.9% |
| Anatomical Diagnosis | 95.9% |
| Total 8 PCI-related key information points | 99.4% |
| Multivessel PCI | 100.0% |
| $\geq 3$ Lesions Treated | 100.0% |
| Bifurcation with $\geq 2$ Stents | 98.3% |
| $\geq 3$ Stents Implanted | 100.0% |
| CTO PCI | 100.0% |
| Total Stent Length >60 mm | 100.0% |
| Complex PCI | 100.0% |
| Current Stents Information | 96.6% |
| CTO = chronic total occlusion. |  |

A qualitative analysis was conducted to understand the reasons for errors made by the framework. As shown in Supplementary Table 4, some inevitable errors arose from the inherent ambiguity and inconsistencies within the CAG section of the reports themselves. However, the PCI sections of the reports were structured using seven key items for each treated lesion—“Location,” “Guiding catheter,” “Guidewire,” “Preballoon,” “Adjuvant balloon,” “DEB,” and “Stent”—which provided a well-organized foundation for data extraction. This pre-existing structure in the free-text report significantly facilitated the conversion into a machine-readable format by the LLM, resulting in high accuracy for extracting PCI-related key points. Supplementary Figure 4 depicts two representative cases from the external test set, showcasing the excellent capability of the framework in converting all the details of a highly complex CAG and PCI case into a machine-readable format.

## DISCUSSION

This study demonstrated that our novel framework, which combines hierarchical annotation with LLMs, can successfully automate the standardization of CAG reports with accuracy comparable to or exceeding that of experienced cardiologists. In both internal and external tests, the framework showed excellent performance in extracting and structuring complex information about coronary anatomy and PCI procedures, achieving up to 99.5% accuracy for CAG-related key points and 98.3% for PCI-related key points. These results suggest that automated standardization of complex medical documents is not only feasible, but can be implemented with high reliability, potentially transforming how we utilize clinical information embedded in unstructured medical reports.

To our knowledge, no previous study has attempted to convert detailed information about coronary anatomy and catheterization procedures from free-text reports into a machine-readable format. A recent systematic review indicated that natural language processing research in the field of cardiology remains underexplored as compared to that in the field of oncology.^25^ Although the availability of coded information in EHRs and claims data has increased substantially, most large-scale observational studies have not utilized detailed information about coronary anatomy or procedural characteristics.^26^ While studies leveraging data from dedicated registries have incorporated such information, this approach requires marked manual effort. Our framework addresses this limitation by automatically converting detailed procedural and anatomical information into a structured, machine-readable format, which promotes several critical capabilities: (1) systematic analysis of large-scale coronary intervention outcomes across different patient populations and institutions, (2) efficient quality assessment and performance monitoring of interventional procedures, (3) facilitation of clinical research by enabling rapid identification of eligible patients for trials based on specific anatomical or procedural criteria,^27^ and (4) providing support for clinical decision-making by making historical procedural data more accessible and analyzable.

The framework developed in this study uses a two-step approach to ensure accuracy and interpretability. In a previous study, while a one-step approach using LLMs was able to capture basic information accurately, it demonstrated difficulty with complex medical reasoning.^9^ For example, in pathology reports, LLMs could accurately identify tumor measurements but had difficulty determining accurate cancer staging. To overcome this limitation, we separated our process into two distinct steps: first, using LLMs to organize the basic information from CAG reports into an expert-defined standardized format, and second, applying specific rules developed by cardiologists to build clinically relevant data. This approach, similar to methods successfully used in pathology research,^10^ combines the LLM’s strength in understanding medical text with cardiologist-designed rules for interpreting nuance and complexity in coronary anatomy and catheterization. Furthermore, an instruction prompt was given to LLM to encode domain-specific knowledge and detailed rules, as guided by cardiologists, to ensure accurate and consistent mapping of the intricate information contained in CAG reports.

The significance of our work extends beyond its immediate performance metrics. By developing and publicly releasing a comprehensive hierarchical annotation schema for CAG reports, we have provided a standardized framework that other institutions can readily adopt or modify for their specific needs. This flexibility is particularly important as healthcare institutions increasingly seek to process sensitive clinical data by using internal systems rather than external services. With the rapid advance in the capabilities of open-source LLMs, our framework offers a practical blueprint for standardization of automated medical documents that could be implemented using various LLM options, while maintaining high accuracy and reliability. The primary advantage is that it saves time and human effort.

### LIMITATIONS

The study had some limitations. First, only CAG reports from hospitals in South Korea were used to validate the robustness of the framework. To ensure thorough validation, the framework should be implemented on CAG reports from more diverse countries. Second, this study utilized only GPT-4o as the LLM for standardization. Nevertheless, our framework was intentionally designed to be model-agnostic and to be able to work with any sufficiently capable LLM, although further testing is needed to determine whether its high accuracy and reliability will be maintained even with less-competent open-source models, such as Llama-3.^28^ Third, unlike the internal validation, the external validation process only evaluated the framework’s accuracy as judged by cardiologists, without direct comparison to cardiologists’ manual reviews. Although the framework achieved excellent accuracy in the external test, a direct comparison with manual reviews would have more clearly demonstrated its capabilities. Fourth, in both the internal and external validation processes, accuracy was evaluated based on only a single output from the LLM. Although we set the LLM’s “temperature”—a parameter that controls the randomness and variability of the model’s responses—to zero, in order to maximize predictability, slight variability remained due to the model’s inherent characteristics. Addressing these limitations in future will help to ensure broader applicability, consistent performance across various LLMs, and even more robust validation methods.

## CONCLUSION

In this study, we developed a novel framework for standardizing free-text CAG reports and extracting complex clinical data. This framework demonstrated excellent accuracy in standardizing CAG reports, indicating its potential for more efficient utilization of detailed clinical data in these reports for cardiovascular research. Future research may explore adaptation of this framework to other types of free-text data.

## Supporting information

STROBE checklist

## Data Availability

All data produced in the present study are available upon reasonable request to the authors.

## Abbreviations

ACC/AHA: American College of Cardiology / American Heart Association
CAG: coronary angiography
CTO: chronic total occlusion
EHR: electronic health records
JSON: JavaScript Object Notation
LLM: large language model
PCI: percutaneous coronary interventions
SYNTAX: Synergy Between Percutaneous Coronary Intervention with Taxus and Cardiac Surgery

## Acknowledgements

This research was supported by a grant of the MD-Phd/Medical Scientist Training Program through the Korea Health Industry Development Institute (KHIDI), funded by the Ministry of Health & Welfare, Republic of Korea.

## Funding Support

This research was supported by a grant of the MD-Phd/Medical Scientist Training Program through the Korea Health Industry Development Institute (KHIDI), funded by the Ministry of Health & Welfare, Republic of Korea, and also by a grant of the Korea Health Technology R&D Project through the Korea Health Industry Development Institute (KHIDI), funded by the Ministry of Health & Welfare, Republic of Korea (grant number: HI22C0452).

## Disclosure of conflicts of interest

SCY reports grants from Daiichi Sankyo. He is a coinventor of granted Korea Patent DP-2023-1223 and DP-2023-0920, and pending Patent Applications DP-2024-0909, DP-2024-0908, DP-2022-1658, DP-2022-1478, and DP-2022-1365 unrelated to current work. SCY is a chief executive officer of PHI Digital Healthcare. Other authors have no potential conflicts of interest to disclose.

## Notes

**Funding information**: This research was supported by a grant of the MD-Phd/Medical Scientist Training Program through the Korea Health Industry Development Institute (KHIDI), funded by the Ministry of Health & Welfare, Republic of Korea.

**Disclosure**: SCY reports grants from Daiichi Sankyo. He is a coinventor of granted Korea Patent DP-2023-1223 and DP-2023-0920, and pending Patent Applications DP-2024-0909, DP-2024-0908, DP-2022-1658, DP-2022-1478, and DP-2022-1365 unrelated to current work. SCY is a chief executive officer of PHI Digital Healthcare. Other authors have no potential conflicts of interest to disclose.

