## Supplementary Appendix for "A Standard Framework for Converting Coronary Angiography Reports into Machine-Readable Format Using Large Language Models"

**Supplemental Appendix**

Song et al.

**Supplementary Method 1. Minimum Reporting Items for Clear Evaluation of Accuracy Reports of Large Language Models in Healthcare (MINI-CLEAR-LLM) checklist^1^**	2

**Supplementary Method 2. Detailed logic of rule-based analysis of machine-readable reports for (A) CAG-related key points and (B) PCI-related key points.**	2

**Supplementary Figure 1. (A) Instruction Prompt and (B) Hierarchical Schema for Standardization of Coronary Angiography (CAG) Reports.**	5

**Supplementary Figure 2. Few-shot Prompt Built From Training Set and Used For In-context Learning in Internal Test.**	13

**Supplementary Figure 3. Examples of free text reports and structured reports from internal test set.**	35

**Supplementary Figure 4. Examples of CAG reports and structured reports from external validation set.**	40

**Supplementary Table 1. Accuracy Comparison for the Analysis on 12 Key Points by the Framework and Two Cardiologists in Internal Test.**	49

**Supplementary Table 2. Qualitative Analysis of Errors (A) the Framework Made and (B) Two Cardiologists Made in Internal Test Set.**	50

**Supplementary Table 3. Accuracy Comparison for the Analysis on 12 Key Points by the Framework and Two Cardiologists in External Test.**	57

**Supplementary Table 4. Qualitative Analysis of Errors the Framework Made in External Test Set.**	58

### **Supplementary Method 1. Minimum Reporting Items for Clear Evaluation of Accuracy Reports of Large Language Models in Healthcare (MINI-CLEAR-LLM) checklist^12^**

In standardization step, GPT-4o (Generative Pretrained Transformer 4 Omni), OpenAI’s recent ChatGPT model released on May 13, 2024, was used.^18^ The temperature, representing the ‘predictability’ of ChatGPT, was set to 0. This forces the model to choose the highest probability option at each step, ensuring consistent, precise, and reliable outputs. For both the internal and external test sets, each CAG report was transformed into a structured format with a single query to ChatGPT. Using the prompt without further adjustments, the queries for the internal test set were conducted on August 19, 2024, and for the external test set on September 4, 2024.

The instruction prompt comprised four components: (1) assigning a role as a data scientist, (2) defining a task to convert free-text report into the machine-readable schema, (3) elaborating rules to adhere and (4) providing domain-specific knowledge. The domain-specific knowledge, which is mainly the numerical segment code of coronary artery location according to the Synergy Between Percutaneous Coronary Intervention with Taxus and Cardiac Surgery (SYNTAX) score, was incorporated to ensure consistent and standardized annotation of lesion and device location.^14^ After presenting the instruction prompt, we included the blank hierarchical annotation schema. During the internal test, we also added a few-shot prompt. Finally, we appended the “unstructured text” annotation, the actual CAG report, and the “structured JSON” annotation, guiding the model to generate the final structured JSON format. Finally, all components—including the instruction prompt, the hierarchical schema, the few-shot prompt, the guiding annotation labeled “unstructured text” and the actual CAG report, as well as the guiding annotation labeled “structured JSON”—were concatenated into a single input for the ChatGPT.

### **Supplementary Method 2. Detailed logic of rule-based analysis of machine-readable reports for (A) CAG-related key points and (B) PCI-related key points.**

**(A) 4 CAG-related Key Points**

1. **Location of Previous Stents**: Simply retrieved from ‘segment_code’ of ‘previous_stents_rearrange’.
2. **Location and Type of Lesion**: First, lesions with a luminal narrowing percentage of 50 or higher, as listed under the 'lesions' section, are selected. For these lesions, segment code was simply retrieved. And the lesion characteristics section is reviewed for type classification. The lesion characteristics were assessed in accordance with the ACC/AHA lesion classification, as initially proposed by Ryan. et al.^15^ Although 'subtotal occlusion' and ‘intra-stent restenosis (ISR)’ are not explicitly listed in the lesion type evaluation criteria, it was judged to be more severe than type B1 but less severe than type C. Given that interventionalists did not document additional characteristics when a lesion was identified as a subtotal occlusion or ISR and considering its clinical severity, it was classified as a type B2 feature. A lesion is classified as type C if it has any type C charateristics; as type B2 if it lacks type C characteristics but has two type B features; as type B1 if it has only one type B characteristic; and as type A if it lacks both type C and type B characteristics.

- Characteristics of Type B: ["tubular", "eccentric", "moderate calcification", "severe calcification", "os", "bifurcation", "irregular"]
- Characteristics of Type B2: [“subtotal occlusion”, “ISR”]
- Characteristics of Type C: ["diffuse", "total occlusion", "CTO"]

3. **Anatomical Diagnosi**s: Anatomical diagnosis (e.g., left main artery disease with 2-vessel disease) was derived from segment code of each lesion in refer to the segment codes of main vessel. Lesions in CAG with lumen narrowing of 50% or more, as well as segments with previous stents regardless of the degree of narrowing, were all taken into consideration.

- SYNTAX segment codes of right coronary artery (RCA) segments: 1, 2, 3, 4, 16, 16a, 16b, 16c
- SYNTAX segment code of left main artery (LM) segments: 5
- SYNTAX segment codes of left anterior descending artery (LAD) segments: 6, 7, 8, 9, 9a, 10, 10a
- SYNTAX segment codes of left circumflex artery (LCx) segments: 11, 12, 12a, 12b, 13, 14, 14a, 14b, 15

4. **Previous Stent Information**: Full information including device name, diameter, length and segment code of previous stents was simply retrieved from ‘previous_stents_rearrange’.

**(B) 8 PCI-related Key Points**

According to Giustino et al.^19^, complex PCI were determined by the presence of one or more of the following criteria in the index procedure: multivessel PCI, implantation of ≥3 stents, treatment of ≥3 lesions, bifurcation PCI using ≥2 stents, total stent length >60 mm, or CTO as target lesion. The six criteria, along with complex PCI, are evaluated. Additionally, full information of ‘current stent’ information including device name, diameter, length and segment code was also included. To evaluate the complexity of a PCI procedure, six criteria are analyzed using the information in ‘rearrange complexity’ in the JSON reports.

1. **Multivessel PCI**: The analysis identifies if lesions are present in the three main coronary arteries, RCA, LAD and LCx. It checks for the presence of segment codes associated with each artery among the segments treated with previous and current stents or drug-eluting balloon (DEB) or thrombectomy by inspecting all segment codes from ‘previous_stents_rearrange’, 'current_stents_rearrange', 'DEB_DCB' and 'thrombus_aspiration'. If segments corresponding to all three arteries (RCA, LAD, and LCx) are found among them, it indicates TRUE.
2. **≥3 Lesions Treated**: It checks for the presence of segment codes treated with current stents or drug-eluting balloon (DEB) or thrombectomy by inspecting all segment codes from 'current_stents_rearrange', 'DEB_DCB' and 'thrombus_aspiration'.If the segments are three or more, it is recored TRUE.
3. **Bifurcation with ≥2 Stents**: Bifurcation junctions, such as the intersections of segments like [“5”, “6”, “11”] which is segment codes of [“LM”, “proximal LAD”, “proximal LCx”] are examined to be present in ‘segment_code’ of ‘previous_stents_rearrange’, 'current_stents_rearrange'. The analysis involves combining the segment codes of both previously implanted stents and currently placed stents. If the combined set of these segments matches any predefined bifurcation junction, it is determined that two stents were deployed at a bifurcation. Every bifurcation junctions represented in SYNTAX segment codes are the following: ["5", "6", "11"], ["6", "7", "9"], ["7", "8", "10"], ["11", "13", "12a"], ["13", "14", "14a"], ["3", "4", "16"], ["13", "14", "15"]
4. **≥3 Stents Implanted**: The number of current stents used during PCI is checked by counting the number of values in ‘length_mm’ of ‘current_stents_rearrange’, and if three or more are found, it is recorded TRUE.
5. **Total Stent Length > 60mm**: The lengths of all current stents (‘length_mm’ of ‘current_stents_rearrange’) are summed, and if the total is 60 or more, it is noted TRUE.
6. **CTO PCI**: The Boolean object ‘CTO’ in ‘pci_details’ is directly referenced if the patient underwent PCI.
7. **Complex PCI**: The six criteria above are reviewed, and if any of these are TRUE, the procedure is classified as complex (TRUE).
8. **Current Stent Information**: Additionally, full information including device name, diameter, length and segment code of current stents was simply retrieved from ‘current_stents_rearrange’.

The Python code of the extraction algorithm is available on <https://github.com/jiuisdisciple/CAGtoJSON>.

### **Supplementary Figure 1. (A) Instruction Prompt and (B) Hierarchical Schema for Standardization of Coronary Angiography (CAG) Reports.**

**(A) Instruction Prompt**

\### Role: You are a knowledgeable data scientist.

\### Task: Convert the last coronary angiography reports into JSON format. Follow the rules and examples. Use JSON structure.

\### Rule:

0. If the patient is atypical, write a concise explanation of why the patient’s CAG report is atypical in the 'check_recommended' object. If the patient is typical, do not include the object. If the report is empty or lacks information appropriate for the JSON format, do not fabricate details and leave the comment as is.

1. **Exclude 'N/A' or Blank Values**: Do not include keys that have a value of 'N/A,' blank (""), or 'null.' Only include keys with a proper value if mentioned. For example, if the PCI-related section is not present, do not add {'pci_detail': []}, but simply omit the object. Additionally, 'false' objects (Boolean type) should not be included. For example, if the patient's lesion is not a CTO (chronic total occlusion), do not write 'CTO': false, but just omit the object.

2. **Segment Code**: The location of the lesion should be converted into a segment code according to the 'Segment definitions.' The object can be either an object or an array. If the lesion is ‘os’ (ostial in location), it should be considered as ‘proximal’ when designating the segment code. Lesions are sequential, meaning that the boundary mentioned in the report implies that all lesions within that boundary are treated. For example, if dLM ~ mLAD were treated, since pLAD is between dLM and mLAD, three segments (LM, pLAD, and mLAD) should be considered as treated. Therefore, location and segment code should be like the following: {'location': ["dLM", "pLAD", "mLAD"], 'segment_code': ["5", "6", "7"] }. Make sure that segment code of RI is '12' and of OM is '12a', which are obviously different. Make sure that 'far dLCx(far distal LCx)' is left PD, whose segment code should be '15'(left PD), rather than '3'(dLCx).

3. **Earlier Previous Stent if Present**: Always place the previous stent array before the lesion array if previous stents are present. If lesions are located within previous stents(ISR, intra-stent restenosis), fill in both the ‘previous_stents’ array and the ‘lesions’ array. Do not omit information from either array.

4. **LN Percentage**: Ignore 'upto' or 'up to' in percentage values. For ‘minimal LN’ (minimal lumen narrowing), assign ‘10’ (10%). For ‘subtotal LN’ (subtotal lumen narrowing), assign ‘95’ (95%). For ‘total occlusion,’ assign ‘100’ (100%).

5. **Lesion Characteristics**: Please ensure that all lesion characteristics are recorded in the "lesion_characteristics" array using only the following terms: "os" (ostial), "ISR" (in-stent restenosis), "NIH" (neointimal hyperplasia), "minimal," "subtotal occlusion," "total occlusion," "discrete," "concentric," "tubular," "eccentric," "irregular," "diffuse," "moderate calcification," "severe calcification," "bifurcation," "milking," and "small caliber." It is especially important to include "os" when it follows the location (e.g., mLAD os, 1st OM os, etc.), as this is frequently missed. Be attentive to accurately capturing and documenting this term. However, if there are additional features not listed above, feel free to include them.

6. **Collateral Flow**: The grades of collateral filling from the contralateral vessel are as follows: Grade 0 = none; Grade I = filling of side branches of the artery to be dilated via collateral channels without visualization of the epicardial segment; Grade II = partial filling of the epicardial segment via collateral channels; Grade III = complete filling of the epicardial segment of the artery being dilated via collateral channels. This should always be written as ‘G’ + ‘grade’ + ‘/III’, but the ‘/III’ should be ignored.

7. **Staged Intervention**: If the report refers to a 2nd PCI or 3rd PCI, mark ‘staged_intervention’ as ‘true.’

8. **PCI Details**: For PCI performed on a chronic total occlusion, mark "CTO" as ‘true.’ If not, do not include the object. All balloons and stents should contain "device," "location," "segment_code," "diameter_mm," and "length_mm."

9. **Guide catheter, microcatheter, guide wire**: Combine all guide catheters used into one array: e.g. ["SAL 1 SH", "XB 6 3.5 SH"]. Combine all microcatheters used into one array: e.g. ["Corsair", "Sasuke", "Caravel"]. Combine all guide wires used into one array: ["Pilot 50", "Runthrough NS"]

9-1. Examples of guide catheter are JR(jurkin right), JL(jurkin left) , AL(amplant left), AR(amplant right) , EBU, XB + catheter size followed (5,6,7,8 Fr) (JL 5-4 means '5Fr JL 4.0 catheter')

9-2. Examples of microcatheter are corsair , sasuke, caravel etc

9-3. Examples of guide wire are {runthrough, sion, sion blue, sion black, pilot, Gaia next 2, BMW, XT-A, Grand slam etc.}

10. **All Balloon Entries**: Ensure that all balloon entries are consolidated into one "all_balloons" array, except for DEB (Drug-Eluted Balloon) or DCB (Drug-Coated Balloon). Kissing balloons are always mentioned in pairs like this:

{Kissing Balloon: NC Balloon 2.5*15 at pLCX~1st OM,

PCI Balloon 2.0*15 at p~dLCX}

Note: The significant space indicates that the "PCI Balloon 2.0*15 at p~dLCX" is also part of the kissing balloon, which is clear to human readers but not to LLMs.

The device can be "PCI balloon," "NC balloon," etc. 'pressure_atm' is often omitted in CAG reports, and when it is, the object should not be filled with length; instead, the object should be omitted from the JSON. Please do not split balloon entries by inserting a stent entry between them.

10-1. **PCI Balloon Examples for 'device' Value**: {Conqueror, Accuforce, FINE PLUS, Genoss, FINEplus, Sapphire, Euphora, SAPPHIRE II PRO, NC Thonic, Selethru, ROVL PLUS, RYUREI, SAVER, LAXA, PANTERA PRO, Tytrak, Pantera LEO, Sapphire NC, NC SELETHRU, Kamui, MINI TREK, Angio Sculpt, MINITREK, Powered3 NC, Pantera NC, Traveler}, followed by {diameter}*{length}.

10-2. **Adjuvant Balloon Examples**: Selethru NC, Accuforce, NC Emerge, NC SAPPHIRE II PRO, Powered NC, Sapphire NC, NC THONIC, NC Euphora, POWERED3NC, PANTERA, PANTERA LEO, NC SELETHRU, Selethru, SAPPHIRE II PRO, SAPPHIRE NC, followed by {diameter}*{length} ({time}).

11. **DEB_DCB entries**: Drug-eluting balloons (a.k.a. drug-coated ballloons) should be separately organized from other balloons.

11-1. **DEB(drug-eluting balloon) examples**: Agent, Prevail, SeQuent, SeQuent Please NEO, GENOSS, followed by {diameter}*{length} ({time})

12. **Stent entries**: Examples are Elunir, Orsiro, BIOFREEDOMultra, BIOFREEDOM, GENOSS, Ultimaster, Synergy, ONYX, Xience skypoint, EluNIR, Xience, SYNERGY MEGATRON, Megatron, CRE8, OSIRO, ULTIMASTER, followed by {diameter}*{length} ({time})

13. **'pressure_atm' in 'all_balloons', 'DEB_DCB' and 'stents'**: If written in range (e.g. 14-20), write the higher value (‘20’ in this case) since it means the interventionalist increased the pressure up to 20.

14. **Drug of FFR and provocation**: Drug, such as adenosine or NTG etc. should be placed in "drug" object.

15. **Thrombus aspiration**: Location, segment code and comment about thrombus aspiration should be noted.

16. **Comment**: Additional context (e.g. "f/u angio", "투여"(sigmart, NTG etc.) and "Rewiring" entries etc.) can be included. If comment is needed, please combine all comments into a single comment. Do not create multiple comment objects, as this can cause confusion during searches.

17.**previous_stents_rearrange**: The previous stents located should be rearranged repeatedly at the end of the JSON format.

18.**current_stents_rearrange**: The current stents placed during this PCI should be rearranged repeatedly at the end of the JSON format.

\### Segment code

SegmentCode	Segment	Abbreviation	Definition

1	RCA proximal	pRCA	From ostium to one half the distance to the acute margin of the heart.

2	RCA mid	mRCA	From end of first segment to acute margin of heart.

3	RCA distal	dRCA	From the acute margin of the heart to the origin of the posterior descending artery.

4	Posterior descending	PD	Running in the posterior interventricular groove.

16	Posterolateral from RCA	PL	Posterolateral branch originating from the distal coronary artery distal to the crux.

16a	Posterolateral from RCA	1st PL	First posterolateral branch from segment 16.

16b	Posterolateral from RCA	2nd PL	Second posterolateral branch from segment 16.

16c	Posterolateral from RCA	3rd PL	Third posterolateral branch from segment 16.

5	Left main	LM	From the ostium of the LCA through bifurcation into left anterior descending and left circumflex branches.

6	LAD proximal	pLAD	Proximal to and including first major septal branch.

7	LAD mid	mLAD	LAD immediately distal to origin of first septal branch and extending to the point where LAD forms an angle (RAO view). If this angle is not identifiable this segment ends at one half the distance from the first septal to the apex of the heart.

8	LAD apical	apical(distal) LAD	Terminal portion of LAD, beginning at the end of previous segment and extending to or beyond the apex.

9	First diagonal	1st Dg or 1st Dx	The first diagonal originating from segment 6 or 7.

9a	First diagonal a	1st Dg or 1st Dx	Additional first diagonal originating from segment 6 or 7, before segment 8.

10	Second diagonal	2nd Dg or 2nd Dx	Second diagonal originating from segment 8 or the transition between segment 7 and 8.

10a	Second diagonal a	2nd Dg or 2nd Dx	Additional second diagonal originating from segment 8.

11	Proximal circumflex	pLCx	Main stem of circumflex from its origin of left main to and including origin of first obtuse marginal branch.

12	Intermediate/anterolateral/ramus intermedius	AL or RI	Branch from trifurcating left main other than proximal LAD or LCX. Belongs to the circumflex territory.

12a	Obtuse marginal 1st OM	a First side branch of circumflex running in general to the area of obtuse margin of the heart.

12b	Obtuse marginal b	2nd OM or 3rd OM	Second or Third additional branch of circumflex running in the same direction as 12.

13	Distal circumflex	dLCx	The stem of the circumflex distal to the origin of the most distal obtuse marginal branch and running along the posterior left atrioventricular grooves. Caliber may be small or artery absent.

14	Left posterolateral	PL	Running to the posterolateral surface of the left ventricle. May be absent or a division of obtuse marginal branch.

14a	Left posterolateral a	2nd PL	Distal from 14 and running in the same direction.

14b	Left posterolateral b	3rd PL Distal from 14 and 14 a and running in the same direction.

15	Posterior descending from LCx	PD or far dLCx	Most distal part of dominant left circumflex when present. Gives origin to septal branches. When this artery is present,

Note) proximal -> p, mid -> m, distal -> d, if there are multiple lesions in same segment, use p, d, m or a, b. For example, of OM has two lesions the lesion must be interpreted as OMa and OMb, which will be 12a, 12b. 3rd obtuse marginal artery should be considered as 12b, since there is no 12c.

LIMA: Left Internal Mammary Artery

RIMA: Right Internal Mammary Artery

VH-IVUS(or VH): virtual histology intravascular ultrasound

**(B) Hierarchical Annotation Schema**

\### JSON format

{

"check_recommended": "string",

"coronary_angiography": {

"catheters": {

"RCA_catheter": ["string"],

"LCA_catheter": ["string"]

},

"previous_stents": [

{

"device": "string",

"location": "string",

"segment_code": ["string"],

"diameter_mm": "number",

"length_mm": "number"

}

],

"lesions": [

{

"lesion_characteristics": ["string"],

"location": "string",

"segment_code": ["string"],

"luminal_narrowing_percentage": "number"

}

],

"collateral_flow": [

{

"grade_out_of_III": "string",

"from_artery": "string",

"to_artery": "string"

}

],

"myocardial_bridging": [

{

"location": "string",

"segment_code": "string",

"comment": ["string"]

}

],

"LIMA": "string",

"RIMA": "string"

},

"staged_intervention": "boolean",

"pci_details": [

{

"CTO": "boolean",

"target_vessel": "string",

"guide_catheter": ["string"],

"microcatheter": ["string"],

"guide_wire": ["string"],

"all_balloons": [

{

"device": "string",

"location": "string",

"segment_code": ["string"],

"diameter_mm": "number",

"length_mm": "number",

"pressure_atm": "number",

"comment": ["string"]

}

],

"DEB_DCB": [

{

"device": "string",

"location": "string",

"segment_code": ["string"],

"diameter_mm": "number",

"length_mm": "number",

"pressure_atm": "number",

"comment": ["string"]

}

],

"thrombus_aspiration": [

{

"location": "string",

"segment_code": ["string"],

"comment": ["string"]

}

],

"stents": [

{

"device": "string",

"location": "string",

"segment_code": ["string"],

"diameter_mm": "number",

"length_mm": "number",

"pressure_atm": "number",

"comment": ["string"]

}

],

"IVUS_used": "boolean",

"OCT_used": "boolean",

"TVC_used": "boolean",

"comments": ["string"]

}

],

"FFR_details": [

{

"location": "string",

"segment_code": ["string"],

"guide_catheter": "string",

"guide_wire": ["string"],

"FFR_baseline": "number",

"drug": "string",

"FFR_hyperemia": "number",

"Pd": "number",

"Pa": "number",

"comment": ["string"]

}

],

"graft_angio": [

{

"catheter": "string",

"location": "string",

"comment": ["string"]

}

],

"peripheral_angio": [

{

"catheter": "string",

"location": "string",

"luminal_narrowing": "number",

"luminal_narrowing_description": "string",

"comment": ["string"]

}

],

"provocation_test": [

{

"location": "string",

"drug": "string",

"stage": "string",

"EKG": "string",

"chest_pain": "string",

"spasm": "string",

"comment": ["string"]

}

],

"contrast_used": {

"type": "string",

"volume_ml": "number"

},

"previous_stents_rearrange": [

{

"device": "string",

"location": "string",

"segment_code": ["string"],

"diameter_mm": "number",

"length_mm": "number"

}

],

"current_stents_rearrange": [

{

"device": "string",

"location": "string",

"segment_code": ["string"],

"diameter_mm": "number",

"length_mm": "number"

}

]

}

### **Supplementary Figure 2. Few-shot Prompt Built From Training Set and Used For In-context Learning in Internal Test.**

The unstructured text on the left and the structured text on the right were all combined in order and input into LLM along with the instruction prompt. (A) Challenging chronic total occlusion (CTO) percutaneous coronary intervention (PCI) scenario. (B) Case with the results of a provocation test. (C) Case with in-stent restenosis (ISR) lesions. (D) Staged PCI case without accompanying coronary angiography results. (E) Representative of common case of CAG and PCI. (F) Case involving an implanted stent with missing device name and length details. (G) Case providing angiography result of coronary artery bypass graft. (H) Case of critical situation with pulseless electrical activity arrest during procedure.

| **(A) CTO PCI scenario** | |
| --- | --- |
| ****Unstructured Text****  RCA c SAL 7-1 SH  Tubular ecc. 30% LN of pRCA  Minimal LN of m-dRCA  Diffuse ecc. 80% LN of PD br  Collateral flow GI/III from RV br to dLAD, PD to septal  LCA c XB 3.5 SH  Tubular ecc. 80% LN of LAD os c severe calcification  Tubular ecc. 90% LN of early Dg os  Chronic Total Occlusion of p~mLAD (TIMI 0)  Diffuse ecc. 50% LN of OM  [PCI at LAD CTO lesion]  Lt: XBG 3.5 SH  Rt: SAL 7-1 SH  -antegrade-  Corsair + 014" G/W : Runthrough → Gaia 1st  Anchoring Balloon : PCI balloon 2.0*15  Sasuke + 014" G/W : Gaia 1st + Gaia 2nd -> Suoh 03  -Retrograde approach-  Corsair 150cm + 014" G/W : Runthrough →Suoh 03(new)  -antegrade-  Corsair + 014"G/W: Gaia 1st → Gaia 3rd  -retrograde-  Corsair 150cm + 014"G/W : Suoh 03  - antegrade -  Corsair + 014" G/W : Gaia 3rd  Anchoring balloon : PCI balloon 2.0*15  Sasuke + 014" G/W : Gaia 3rd + Suoh 03→ Gaia 3rd + Sion  Caravel +014" G/W : Suoh 03  - antegrade-  Corsair + 014"G/W: Gaia 3rd → Gaia 2nd → Gaia 3rd -> Gaia1st  Balloon : PCI balloon 2.0*15 at p-mLAD  IVUS (+)  Stent : Biomime 3.0*40(9atm) at p-mLAD  IVUS (+)  Stent : Firehawk 3.5*29 (16atm) at pLAD - stent overlapping  f/u angio : R/S > 10% &amp; sent underexpansion  Adjuvant balloon : NC baloon 3..0*15 (20atm) at p-mLAD  IVUS (+)  f/u angio : No R/S    Contrast : Visipaque 500cc | ****Structured JSON****  ```json  {  "coronary_angiography": {  "catheters": {  "RCA_catheter": ["SAL 7-1 SH"],  "LCA_catheter": ["XB 3.5 SH"]  },  "lesions": [  {  "lesion_characteristics": ["tubular", "eccentric"],  "location": "pRCA",  "segment_code": ["1"],  "luminal_narrowing_percentage": 30  },  {  "lesion_characteristics": ["minimal"],  "location": "m-dRCA",  "segment_code": ["2", "3"],  "luminal_narrowing_percentage": 10  },  {  "lesion_characteristics": ["diffuse", "eccentric"],  "location": "PD",  "segment_code": ["4"],  "luminal_narrowing_percentage": 80  },  {  "lesion_characteristics": ["tubular", "eccentric", "os", "severe calcification"],  "location": "LAD os",  "segment_code": ["6"],  "luminal_narrowing_percentage": 80  },  {  "lesion_characteristics": ["tubular", "eccentric"],  "location": "early Dg os",  "segment_code": ["9"],  "luminal_narrowing_percentage": 90  },  {  "lesion_characteristics": ["total occlusion"],  "location": "p~mLAD",  "segment_code": ["6", "7"],  "luminal_narrowing_percentage": 100  },  {  "lesion_characteristics": ["diffuse", "eccentric"],  "location": "OM",  "segment_code": ["12a"],  "luminal_narrowing_percentage": 50  }  ],  "collateral_flow": [  {  "grade_out_of_III": "I",  "from_artery": "RV br",  "to_artery": "dLAD"  },  {  "grade_out_of_III": "I",  "from_artery": "PD",  "to_artery": "septal"  }  ]  },  "pci_details": [  {  "CTO": true,  "target_vessel": "LAD",  "guide_catheter": ["XBG 3.5 SH", "SAL 7-1 SH"],  "microcatheter": ["Corsair", "Sasuke", "Caravel"],  "guide_wire": ["Runthrough", "Gaia 1st", "Gaia 2nd", "Suoh 03", "Gaia 3rd", "Sion"],  "all_balloons": [  {  "device": "PCI balloon",  "location": "p-mLAD",  "segment_code": ["6", "7"],  "diameter_mm": 2.0,  "length_mm": 15  },  {  "device": "NC balloon",  "location": "p-mLAD",  "segment_code": ["6", "7"],  "diameter_mm": 3.0,  "length_mm": 15,  "pressure_atm": 20  }  ],  "stents": [  {  "device": "Biomime",  "location": "p-mLAD",  "segment_code": ["6", "7"],  "diameter_mm": 3.0,  "length_mm": 40,  "pressure_atm": 9  },  {  "device": "Firehawk",  "location": "pLAD",  "segment_code": ["6"],  "diameter_mm": 3.5,  "length_mm": 29,  "pressure_atm": 16,  "comment": ["stent overlapping"]  }  ],  "IVUS_used": true,  "comments": ["f/u angio: R/S > 10% & stent underexpansion", "f/u angio: No R/S"]  }  ],  "contrast_used": {  "type": "Visipaque",  "volume_ml": 500  },  "current_stents_rearrange": [  {  "device": "Biomime",  "location": "p-mLAD",  "segment_code": ["6", "7"],  "diameter_mm": 3.0,  "length_mm": 40  },  {  "device": "Firehawk",  "location": "pLAD",  "segment_code": ["6"],  "diameter_mm": 3.5,  "length_mm": 29  }  ]  }  ``` |
| **(B) Case with Provocation Test** | |
| ****Unstructured Text****  LCA c JL 5-4  No significant LN  RCA c JR 5-4 → AR 1  No significant LN  [Provocation test]  1.RCA  Stage III : EKG(-), Chest pain(-), Spasm(-)  2. LCA  Stage III : EKG(-), Chest pain(-), Spasm(-)  → NTG injection via I.C  Contrast : Scanlux 150cc | ****Structured JSON****  ```json  {  "coronary_angiography": {  "catheters": {  "RCA_catheter": ["JR 5-4", "AR 1"],  "LCA_catheter": ["JL 5-4"]  }  },  "provocation_test": [  {  "location": "RCA",  "drug": "NTG",  "stage": "Stage III",  "EKG": "negative",  "chest_pain": "negative",  "spasm": "negative"  },  {  "location": "LCA",  "drug": "NTG",  "stage": "Stage III",  "EKG": "negative",  "chest_pain": "negative",  "spasm": "negative"  }  ],  "contrast_used": {  "type": "Scanlux",  "volume_ml": 150  }  }  ``` |
| **(C) Case with multiple in-stent restenosis lesions.** | |
| ****Unstructured Text****  RCA c JR 5-4  Tubular ecc. 60% LN of p-mRCA  Minimal LN of dRCA  LCA c JL 5-4  ISR 100% previous stent at LM-pLAD(cypher 3.5*33), pLAD(cypher 3.5*18), mLAD(cypher 3.0*28) [2007.01.31] → pLAD(taxus 3.5*20)[2007.08.22]  Tubular ecc. 80% LN of LCx os  Tubular ecc. 50% LN of dLCx  Graft angio c JL  LIMA to dLAD    [PCI at LCx os]  SPB 6-3.5 SH → SPB 6-3 SH → JLG 6-4 SH → JLG 6-3.5 SH  014"G/W: Sion blue ES  IVUS (+)  Balloon : PCI balloon 2.5*15(6atm) at  Stent: Biomime3.5*24(10atm) at LM-pLCx  f/u angio: 10% < R/S &amp; stent under expansion  adjuvant: NC balloon 3.5*13(14atm)  f/u angio: No R/S  [FFR at dLCx]  JLG 6-3.5 SH  014"pressure wire  baseline : 0.95  adenosine (IV) : 0.87 87/100mmHg    P-angio c JR  No sig. LN at Lt. CFA  contrast: Xenentix 350cc | ****Structured JSON****  ```json  {  "coronary_angiography": {  "catheters": {  "RCA_catheter": ["JR 5-4"],  "LCA_catheter": ["JL 5-4"]  },  "previous_stents": [  {  "device": "cypher",  "location": "LM-pLAD",  "segment_code": ["5", "6"],  "diameter_mm": 3.5,  "length_mm": 33  },  {  "device": "cypher",  "location": "pLAD",  "segment_code": ["6"],  "diameter_mm": 3.5,  "length_mm": 18  },  {  "device": "cypher",  "location": "mLAD",  "segment_code": ["7"],  "diameter_mm": 3.0,  "length_mm": 28  },  {  "device": "taxus",  "location": "pLAD",  "segment_code": ["6"],  "diameter_mm": 3.5,  "length_mm": 20  }  ],  "lesions": [  {  "lesion_characteristics": ["tubular", "eccentric"],  "location": "p-mRCA",  "segment_code": ["1", "2"],  "luminal_narrowing_percentage": 60  },  {  "lesion_characteristics": ["minimal"],  "location": "dRCA",  "segment_code": ["3"],  "luminal_narrowing_percentage": 10  },  {  "lesion_characteristics": ["ISR", "total occlusion"],  "location": "LM-pLAD",  "segment_code": ["5", "6"],  "luminal_narrowing_percentage": 100  },  {  "lesion_characteristics": ["ISR", "total occlusion"],  "location": "pLAD",  "segment_code": ["6"],  "luminal_narrowing_percentage": 100  },  {  "lesion_characteristics": ["ISR", "total occlusion"],  "location": "mLAD",  "segment_code": ["7"],  "luminal_narrowing_percentage": 100  },  {  "lesion_characteristics": ["tubular", "eccentric"],  "location": "LCx os",  "segment_code": ["11"],  "luminal_narrowing_percentage": 80  },  {  "lesion_characteristics": ["tubular", "eccentric"],  "location": "dLCx",  "segment_code": ["13"],  "luminal_narrowing_percentage": 50  }  ],  "graft_angio": [  {  "catheter": "JL",  "location": "LIMA to dLAD"  }  ]  },  "pci_details": [  {  "target_vessel": "LCx os",  "guide_catheter": ["SPB 6-3.5 SH", "SPB 6-3 SH", "JLG 6-4 SH", "JLG 6-3.5 SH"],  "guide_wire": ["Sion blue ES"],  "IVUS_used": true,  "all_balloons": [  {  "device": "PCI balloon",  "location": "LCx os",  "segment_code": ["11"],  "diameter_mm": 2.5,  "length_mm": 15,  "pressure_atm": 6  },  {  "device": "NC balloon",  "location": "LCx os",  "segment_code": ["11"],  "diameter_mm": 3.5,  "length_mm": 13,  "pressure_atm": 14  }  ],  "stents": [  {  "device": "Biomime",  "location": "LM-pLCx",  "segment_code": ["5", "11"],  "diameter_mm": 3.5,  "length_mm": 24,  "pressure_atm": 10,  "comment": ["f/u angio: 10% < R/S & stent under expansion"]  }  ],  "comments": ["f/u angio: No R/S"]  }  ],  "FFR_details": [  {  "location": "dLCx",  "segment_code": ["13"],  "guide_catheter": "JLG 6-3.5 SH",  "guide_wire": ["014\" pressure wire"],  "FFR_baseline": 0.95,  "drug": "adenosine (IV)",  "FFR_hyperemia": 0.87,  "Pd": 87,  "Pa": 100  }  ],  "peripheral_angio": [  {  "catheter": "JR",  "location": "Lt. CFA",  "luminal_narrowing": 0,  "luminal_narrowing_description": "No sig. LN"  }  ],  "contrast_used": {  "type": "Xenentix",  "volume_ml": 350  },  "previous_stents_rearrange": [  {  "device": "cypher",  "location": "LM-pLAD",  "segment_code": ["5", "6"],  "diameter_mm": 3.5,  "length_mm": 33  },  {  "device": "cypher",  "location": "pLAD",  "segment_code": ["6"],  "diameter_mm": 3.5,  "length_mm": 18  },  {  "device": "cypher",  "location": "mLAD",  "segment_code": ["7"],  "diameter_mm": 3.0,  "length_mm": 28  },  {  "device": "taxus",  "location": "pLAD",  "segment_code": ["6"],  "diameter_mm": 3.5,  "length_mm": 20  }  ],  "current_stents_rearrange": [  {  "device": "Biomime",  "location": "LM-pLCx",  "segment_code": ["5", "11"],  "diameter_mm": 3.5,  "length_mm": 24  }  ]  }  ``` |
| **(D) Staged PCI case without accompanying CAG results.** | |
| ****Unstructured Text****  ===== 2nd PCI =====  [PCI at RCA]  SAL 7-1 SH  buddy 014" G/W : bmw + pilot  balloon: PCI balloon 2.0*15  stent: synergy 2.5*28 (16atm) at dRCA~PL  synergy 3.0*38 (11atm) at m~dRCA overlap (+)  IVUS was done  f/u angio : 10%> R/S  adjuvant balloon: NC 3.5*13 (16atm)  IVUS was done  f/u angio : No R/S  contrast :scanlux 230cc | **Structured JSON**  ```json  {  "staged_intervention": true,  "pci_details": [  {  "target_vessel": "RCA",  "guide_catheter": ["SAL 7-1 SH"],  "guide_wire": ["BMW", "Pilot"],  "all_balloons": [  {  "device": "PCI balloon",  "location": "dRCA",  "segment_code": ["3"],  "diameter_mm": 2.0,  "length_mm": 15  },  {  "device": "NC balloon",  "location": "mRCA",  "segment_code": ["2"],  "diameter_mm": 3.5,  "length_mm": 13,  "pressure_atm": 16  }  ],  "stents": [  {  "device": "Synergy",  "location": "dRCA~PL",  "segment_code": ["3", "16"],  "diameter_mm": 2.5,  "length_mm": 28,  "pressure_atm": 16  },  {  "device": "Synergy",  "location": "m~dRCA",  "segment_code": ["2", "3"],  "diameter_mm": 3.0,  "length_mm": 38,  "pressure_atm": 11,  "comment": ["overlap"]  }  ],  "IVUS_used": true,  "comments": ["f/u angio: 10%> R/S", "f/u angio: No R/S"]  }  ],  "contrast_used": {  "type": "scanlux",  "volume_ml": 230  },  "current_stents_rearrange": [  {  "device": "Synergy",  "location": "dRCA~PL",  "segment_code": ["3", "16"],  "diameter_mm": 2.5,  "length_mm": 28  },  {  "device": "Synergy",  "location": "m~dRCA",  "segment_code": ["2", "3"],  "diameter_mm": 3.0,  "length_mm": 38  }  ]  }  ``` |
| **(E) Representative of common case of CAG and PCI.** | |
| ****Unstructured Text****  RCA : JR 5*4  No sig. LN  LCA : JL 7*3.5(S.H)  60% Tubular eccentric LN of Lt.Main ostial  95% Tubular eccentric LN of p LAD( ISR at p LAD:Taxus 3.0*24)  < PCI at LAD - Lt.MAIN >  JLG 7*3.5(S.H)  014 " G/W (Runthrough)  -> IVUS : from Previous stent to Lt.MAIN os  -> PTCA balloon ( Cutting 3.0*10 : p LAD) : 10atm-10sec  -> Coronary stent ( Endeavor resolute 3.5*30) : 16atm-10sec  -> IVUS  contrast : Visipaque 110 ml | ****Structured JSON****  ```json  {  "coronary_angiography": {  "catheters": {  "RCA_catheter": ["JR 5*4"],  "LCA_catheter": ["JL 7*3.5(S.H)"]  },  "previous_stents": [  {  "device": "Taxus",  "location": "pLAD",  "segment_code": ["6"],  "diameter_mm": 3.0,  "length_mm": 24  }  ],  "lesions": [  {  "lesion_characteristics": ["tubular", "eccentric"],  "location": "LM",  "segment_code": ["5"],  "luminal_narrowing_percentage": 60  },  {  "lesion_characteristics": ["tubular", "eccentric", "ISR"],  "location": "pLAD",  "segment_code": ["6"],  "luminal_narrowing_percentage": 95  }  ]  },  "pci_details": [  {  "target_vessel": "LAD - Lt.MAIN",  "guide_catheter": ["JLG 7*3.5(S.H)"],  "guide_wire": ["Runthrough"],  "all_balloons": [  {  "device": "Cutting",  "location": "pLAD",  "segment_code": ["6"],  "diameter_mm": 3.0,  "length_mm": 10,  "pressure_atm": 10  }  ],  "stents": [  {  "device": "Endeavor resolute",  "location": "pLAD",  "segment_code": ["6"],  "diameter_mm": 3.5,  "length_mm": 30,  "pressure_atm": 16  }  ],  "IVUS_used": true  }  ],  "contrast_used": {  "type": "Visipaque",  "volume_ml": 110  },  "previous_stents_rearrange": [  {  "device": "Taxus",  "location": "pLAD",  "segment_code": ["6"],  "diameter_mm": 3.0,  "length_mm": 24  }  ],  "current_stents_rearrange": [  {  "device": "Endeavor resolute",  "location": "pLAD",  "segment_code": ["6"],  "diameter_mm": 3.5,  "length_mm": 30  }  ]  }  ``` |
| **(F) Case involving an implanted stent with missing information.** | |
| ****Unstructured Text****  RCA c JR 5-4  Tubular ecc. 30% LN of p~mRCA  LCA c JL 5-4  Tubular ecc. 40% LN of pLAD  Patent previous stent at mLAD(unknown stent(2002)  Tubular ecc. 30% LN of pLCx  Diffuse irr. 40% LN of OM  Tubular ecc. 40% LN of dLCx  Contrast : Visipaque 80cc | ****Structured JSON****  ```json  {  "coronary_angiography": {  "catheters": {  "RCA_catheter": ["JR 5-4"],  "LCA_catheter": ["JL 5-4"]  },  "previous_stents": [  {  "device": "Unknown",  "location": "mLAD",  "segment_code": ["7"],  "diameter_mm": null,  "length_mm": null,  "comment": ["2002"]  }  ],  "lesions": [  {  "lesion_characteristics": ["tubular", "eccentric"],  "location": "p~mRCA",  "segment_code": ["1". "2"],  "luminal_narrowing_percentage": 30  },  {  "lesion_characteristics": ["tubular", "eccentric"],  "location": "pLAD",  "segment_code": ["6"],  "luminal_narrowing_percentage": 40  },  {  "lesion_characteristics": ["tubular", "eccentric"],  "location": "pLCx",  "segment_code": ["11"],  "luminal_narrowing_percentage": 30  },  {  "lesion_characteristics": ["diffuse", "irregular"],  "location": "OM",  "segment_code": ["12a"],  "luminal_narrowing_percentage": 40  },  {  "lesion_characteristics": ["tubular", "eccentric"],  "location": "dLCx",  "segment_code": ["13"],  "luminal_narrowing_percentage": 40  }  ]  },  "contrast_used": {  "type": "Visipaque",  "volume_ml": 80  },  "previous_stents_rearrange": [  {  "device": "Unknown",  "location": "mLAD",  "segment_code": ["7"],  "diameter_mm": null,  "length_mm": null,  "comment": ["2002"]  }  ]  }  ``` |
| **(G) Case providing angiography result of coronary artery bypass graft.** | |
| ****Unstructured Text****  Graft angio 1: JR 5-4 → AL 5-1  30% NIH at Aorta to dRCA c SVG(Biofreedom 2.5*24)  Tubular irr 80% LN of Aorta to dRCA c mSVG  Graft angio 2: AL 5-1  Patent at Aorta to 1st OM c pSVG (Desyne 3.5*18)  50% ISR at Aorta to 1st OM c mSVG(Nobori 2.5*14)  Graft angio 3: JR 5-4→ IM  Patent LIMA to dLAD  visipaque: 130cc | ****Structured JSON****  ```json  {  "check_recommended": "This coronary angiography report only provides information on the graft angiography, possibly due to prior CABG. It is recommended that the clinician review this.",  "graft_angio": [  {  "catheter": "JR 5-4",  "location": "Aorta to dRCA",  "comment": ["30% NIH at Aorta to dRCA c SVG(Biofreedom 2.5*24)", "Tubular irr 80% LN of Aorta to dRCA c mSVG"]  },  {  "catheter": "AL 5-1",  "location": "Aorta to 1st OM",  "comment": ["Patent at Aorta to 1st OM c pSVG (Desyne 3.5*18)", "50% ISR at Aorta to 1st OM c mSVG(Nobori 2.5*14)"]  },  {  "catheter": "JR 5-4",  "location": "LIMA to dLAD",  "comment": ["Patent LIMA to dLAD"]  }  ],  "contrast_used": {  "type": "visipaque",  "volume_ml": 130  }  }  ``` |
| **(H) Case of emergency situation.** | |
| ****Unstructured Text****  LCA with JL 5-4 revealed  No significant LN  Collateral flow from dLCx to dRCA  RCA with JR 5-4 revealed  Total occlusion of mRCA(TIMI 0)  [PCI at RCA]  Rt.CFV → 6F sheath → Temporary pacemaker  014 inch guidewire : runthrough  Guiding catheter : JR 7-4 side hole  Preballoon : Lacrosse 2.5x15  F/U angio : No improvement  Thrombus aspiration 6F → Thrombus aspiation not well done → 7F change  Red thrombi and white thrombi was removed.  F/U angio : No improvement with intraluminal filling defect  Preballoon : Lacrosse 2.5x15  F/U angio : No interval change  Thrombus aspiration with 7F catheter  →Red thrombi  Attempted thrombus suction using the mother and child method with inner guiding, but could not reach the thrombus location because the guiding length and inner guiding length were almost the same.  Preballoon : Euphora 3.5x15  F/U angio : Total occlusion with TIMI 1 flow  →BP down, dyspnea, Mental change  Stent : Orsiro 3.0x40  F/U angio : no residual stenosis with improved distal flow  ***PEA arrest  Intubation, VA ECMO insertion were done. (Rt. CFA, Lt. CFV)  Bedside TTE: large amount of pericardial effusion  →Pericardiocentesis was done. 1700cc blood was drained. | ****Structured JSON****  ```json  {  "check_recommended": "Despite stent placement improving flow, the patient suffered PEA arrest, requiring intubation and VA ECMO. A large pericardial effusion was identified and drained.",  "coronary_angiography": {  "catheters": {  "RCA_catheter": ["JR 5-4"],  "LCA_catheter": ["JL 5-4"]  },  "lesions": [  {  "lesion_characteristics": ["total occlusion"],  "location": "mRCA",  "segment_code": ["2"],  "luminal_narrowing_percentage": 100  }  ],  "collateral_flow": [  {  "from_artery": "dLCx",  "to_artery": "dRCA"  }  ]  },  "pci_details": [  {  "target_vessel": "RCA",  "guide_catheter": ["JR 7-4 side hole"],  "guide_wire": ["runthrough"],  "all_balloons": [  {  "device": "Lacrosse",  "location": "mRCA",  "segment_code": ["2"],  "diameter_mm": 2.5,  "length_mm": 15  },  {  "device": "Lacrosse",  "location": "mRCA",  "segment_code": ["2"],  "diameter_mm": 2.5,  "length_mm": 15  },  {  "device": "Euphora",  "location": "mRCA",  "segment_code": ["2"],  "diameter_mm": 3.5,  "length_mm": 15  }  ],  "thrombus_aspiration": [  {  "location": "mRCA",  "segment_code": ["2"],  "comment": ["Thrombus aspiration with 6F and 7F catheters", "Red thrombi and white thrombi removed", "Mother and child method used for thrombus suction"]  }  ],  "stents": [  {  "device": "Orsiro",  "location": "mRCA",  "segment_code": ["2"],  "diameter_mm": 3.0,  "length_mm": 40  }  ],  "comments": ["F/U angio: no residual stenosis with improved distal flow", "PEA arrest occurred", "Intubation, VA ECMO insertion performed", "Bedside TTE showed large amount of pericardial effusion", "Pericardiocentesis performed, approximately 1700cc blood drained"]  }  ],  "previous_stents_rearrange": [],  "current_stents_rearrange": [  {  "device": "Orsiro",  "location": "mRCA",  "segment_code": ["2"],  "diameter_mm": 3.0,  "length_mm": 40  }  ]  }  ``` |

### **Supplementary Figure 3. Examples of free text reports and structured reports from internal test set.**

| **(A) Complex case containing compact information about lesions. Two cardiologists made mistakes in identifying location and type of lesions, while LLM made no mistakes.** | |
| --- | --- |
| ****Unstructured Text****  RCA c JR 5-4  Tubular ecc. 30% LN of pRCA  Tubular ecc. 60% LN of mRCA  Tubular ecc. 50% LN of PD os  LCA c JL 5-4  Diffuse ecc. 50% LN of pLAD  Tubular irr. 50% LN of Early Dg  Patent previous stent at mLAD(Cypher 2.5*23)  Tubular ecc. 40% LN of pLCX  50% ISR at far dLCx (Cypher 2.5*23)  Contrast : Scanlux 60cc | ****Structured Text****  ```json  {  "coronary_angiography": {  "catheters": {  "RCA_catheter": ["JR 5-4"],  "LCA_catheter": ["JL 5-4"]  },  "previous_stents": [  {  "device": "Cypher",  "location": "mLAD",  "segment_code": ["7"],  "diameter_mm": 2.5,  "length_mm": 23  },  {  "device": "Cypher",  "location": "far dLCx",  "segment_code": ["13"],  "diameter_mm": 2.5,  "length_mm": 23  }  ],  "lesions": [  {  "lesion_characteristics": ["tubular", "eccentric"],  "location": "pRCA",  "segment_code": ["1"],  "luminal_narrowing_percentage": 30  },  {  "lesion_characteristics": ["tubular", "eccentric"],  "location": "mRCA",  "segment_code": ["2"],  "luminal_narrowing_percentage": 60  },  {  "lesion_characteristics": ["tubular", "eccentric", "os"],  "location": "PD os",  "segment_code": ["4"],  "luminal_narrowing_percentage": 50  },  {  "lesion_characteristics": ["diffuse", "eccentric"],  "location": "pLAD",  "segment_code": ["6"],  "luminal_narrowing_percentage": 50  },  {  "lesion_characteristics": ["tubular", "irregular"],  "location": "Early Dg",  "segment_code": ["9"],  "luminal_narrowing_percentage": 50  },  {  "lesion_characteristics": ["tubular", "eccentric"],  "location": "pLCx",  "segment_code": ["11"],  "luminal_narrowing_percentage": 40  },  {  "lesion_characteristics": ["ISR"],  "location": "far dLCx",  "segment_code": ["13"],  "luminal_narrowing_percentage": 50  }  ]  },  "contrast_used": {  "type": "Scanlux",  "volume_ml": 60  },  "previous_stents_rearrange": [  {  "device": "Cypher",  "location": "mLAD",  "segment_code": ["7"],  "diameter_mm": 2.5,  "length_mm": 23  },  {  "device": "Cypher",  "location": "far dLCx",  "segment_code": ["13"],  "diameter_mm": 2.5,  "length_mm": 23  }  ]  }  ``` |
| **(B) Case of which fractional flow reserve (FFR) information was well identified in JSON report.** | |
| ****Unstructured Text****  RCA c JR 5-4  minimal LN of p-mRCA  LCA c JL 5-4  patent previous stent at pLAD (Biomatrix Flex 4*14)  tubular ecc. 70% LN of mLAD  [FFR at LAD]  XB 6 3.5 SH  pressure wire  base0.92  sigmart IC 0.69(58/86mmHg)  [PCI at LAD]  014" G/W  balloon: 2.5*15  stent: resolute onyx 4.0*22 (14atm) at m-dLAD  sigmart via IC  f/u angio: 10%>R/S, underexpansion  adjuvant : NC 3.5*8 (20atm)  TVC was done  f/u angio : No R/S  contrast : visipaque 300ml | ****Structured JSON****  ```json  {  "coronary_angiography": {  "catheters": {  "RCA_catheter": ["JR 5-4"],  "LCA_catheter": ["JL 5-4"]  },  "previous_stents": [  {  "device": "Biomatrix Flex",  "location": "pLAD",  "segment_code": ["6"],  "diameter_mm": 4.0,  "length_mm": 14  }  ],  "lesions": [  {  "lesion_characteristics": ["minimal"],  "location": "p-mRCA",  "segment_code": ["1", "2"],  "luminal_narrowing_percentage": 10  },  {  "lesion_characteristics": ["tubular", "eccentric"],  "location": "mLAD",  "segment_code": ["7"],  "luminal_narrowing_percentage": 70  }  ]  },  "FFR_details": [  {  "location": "LAD",  "segment_code": ["6", "7"],  "guide_catheter": "XB 6 3.5 SH",  "guide_wire": ["pressure wire"],  "FFR_baseline": 0.92,  "drug": "sigmart IC",  "FFR_hyperemia": 0.69,  "Pd": 58,  "Pa": 86  }  ],  "pci_details": [  {  "target_vessel": "LAD",  "guide_catheter": ["XB 6 3.5 SH"],  "guide_wire": ["014\" G/W"],  "all_balloons": [  {  "device": "PCI balloon",  "location": "m-dLAD",  "segment_code": ["7", "8"],  "diameter_mm": 2.5,  "length_mm": 15  },  {  "device": "NC balloon",  "location": "m-dLAD",  "segment_code": ["7", "8"],  "diameter_mm": 3.5,  "length_mm": 8,  "pressure_atm": 20  }  ],  "stents": [  {  "device": "resolute onyx",  "location": "m-dLAD",  "segment_code": ["7", "8"],  "diameter_mm": 4.0,  "length_mm": 22,  "pressure_atm": 14  }  ],  "TVC_used": true,  "comments": ["f/u angio: 10%>R/S, underexpansion", "f/u angio: No R/S"]  }  ],  "contrast_used": {  "type": "visipaque",  "volume_ml": 300  },  "previous_stents_rearrange": [  {  "device": "Biomatrix Flex",  "location": "pLAD",  "segment_code": ["6"],  "diameter_mm": 4.0,  "length_mm": 14  }  ],  "current_stents_rearrange": [  {  "device": "resolute onyx",  "location": "m-dLAD",  "segment_code": ["7", "8"],  "diameter_mm": 4.0,  "length_mm": 22  }  ]  }  ``` |

### **Supplementary Figure 4. Examples of CAG reports and structured reports from external validation set.**

| **(A) Case whose JSON report captured information about 5 previous stents and 1 current stent perfectly. This complicated case was highly complex PCI, being ‘multivessel PCI’, ‘three or more lesions treated’ PCI and ‘bifurcation lesion treated with two or more stents’ in one case.** | |
| --- | --- |
| ****Unstructured Text****  *Date *Date of CAG PCI*:  *Coronary Angiography*  [**Findings**  Lt RA c/ 6Fr sheath  RCA c/ JR  ISR prev. pRCA stent (taxus 3.5x20) [06.4.17]  mild ISR prev. mRCA stent (taxus 3.5x28) [06.4.17]  LCA c/ JL  subtotal occlusion of LAD os  patent prev. mLAD stent (taxus 3.5x32) [06.4.17]  patent prev. 1st Dg stent (taxus 2.75x32) [06.4.17]  s/p PCI of LAD/RCA (****hospital,2010.08)  patent prev. dLCx stent (xience 3.0x33) [12.7.26]    PCI of LAD os  IVUS guided PCI  preballoon 3.5x15  overlapped stent prev. mLAD stent to LM  exderexpansion : adjuvant balloon 4.0x10 for LM  PCI of RCA  balloon was not delivered  Guideliner backup  preballoon 3.5x15  DCB  Conclusion:**C-MODE [24.1.16]  CAOD  PCI of mLAD (taxus 3.5x32), Dg (2.75x32) [06.4.17]  s/p PCI of LAD/RCA (****hospital,2010.08)  PCI of dLCx (xience 3.0x33) [12.7.26]  s/p PCI of LM to pLAD (Onxy 3.5x18), DCB of mRCA (Prevail 3.5x20) [24.1.16]  Rt thalamic infarction  HTN  dm by local  s/p DU bleeding  current smoker]  *PCI*  [  Diagnosis before procedure:Stable angina  **Target lesion 1**{  location:LM to LAD  Guiding catheter:JL 6F-3.5  Guidewire:Asahi sion blue, Asahi sion  Preballoon:Selethru NC 3.5*15  Adjuvant balloon:Selethru NC 4.5*10 (10:34)  DEB:  Stent:Onyx 3.5*18 (10:25)}  **Target lesion 2**{  location:mRCA  Guiding catheter:JR  Guidewire:sion  Preballoon:Selethru NC 3.5*15  Adjuvant balloon:  DEB:Prevail 3.5x20  Stent:}  **Target lesion 3**{  location:  Guiding catheter:  Guidewire:  Preballoon:  Adjuvant balloon:  DEB:  Stent:}  **Target lesion 4**{  location:  Guiding catheter:  Guidewire:  Preballoon:  Adjuvant balloon:  DEB:  Stent:}** Diagnosis after procedure:  **Stable angina] | ****Structured JSON****  `` ```json  {  "coronary_angiography": {  "catheters": {  "RCA_catheter": ["JR"],  "LCA_catheter": ["JL"]  },  "previous_stents": [  {  "device": "taxus",  "location": "pRCA",  "segment_code": ["1"],  "diameter_mm": 3.5,  "length_mm": 20  },  {  "device": "taxus",  "location": "mRCA",  "segment_code": ["2"],  "diameter_mm": 3.5,  "length_mm": 28  },  {  "device": "taxus",  "location": "mLAD",  "segment_code": ["7"],  "diameter_mm": 3.5,  "length_mm": 32  },  {  "device": "taxus",  "location": "1st Dg",  "segment_code": ["9"],  "diameter_mm": 2.75,  "length_mm": 32  },  {  "device": "xience",  "location": "dLCx",  "segment_code": ["13"],  "diameter_mm": 3.0,  "length_mm": 33  }  ],  "lesions": [  {  "lesion_characteristics": ["subtotal occlusion", "os"],  "location": "LAD os",  "segment_code": ["6"],  "luminal_narrowing_percentage": 95  }  ]  },  "pci_details": [  {  "target_vessel": "LM to LAD",  "guide_catheter": ["JL 6F-3.5"],  "guide_wire": ["Asahi sion blue", "Asahi sion"],  "all_balloons": [  {  "device": "Selethru NC",  "location": "LM to LAD",  "segment_code": ["5", "6", "7"],  "diameter_mm": 3.5,  "length_mm": 15  },  {  "device": "Selethru NC",  "location": "LM",  "segment_code": ["5"],  "diameter_mm": 4.5,  "length_mm": 10,  "pressure_atm": 10  }  ],  "stents": [  {  "device": "Onyx",  "location": "LM to pLAD",  "segment_code": ["5", "6"],  "diameter_mm": 3.5,  "length_mm": 18,  "pressure_atm": 25  }  ]  },  {  "target_vessel": "mRCA",  "guide_catheter": ["JR"],  "guide_wire": ["sion"],  "all_balloons": [  {  "device": "Selethru NC",  "location": "mRCA",  "segment_code": ["2"],  "diameter_mm": 3.5,  "length_mm": 15  }  ],  "DEB_DCB": [  {  "device": "Prevail",  "location": "mRCA",  "segment_code": ["2"],  "diameter_mm": 3.5,  "length_mm": 20  }  ]  }  ],  "previous_stents_rearrange": [  {  "device": "taxus",  "location": "pRCA",  "segment_code": ["1"],  "diameter_mm": 3.5,  "length_mm": 20  },  {  "device": "taxus",  "location": "mRCA",  "segment_code": ["2"],  "diameter_mm": 3.5,  "length_mm": 28  },  {  "device": "taxus",  "location": "mLAD",  "segment_code": ["7"],  "diameter_mm": 3.5,  "length_mm": 32  },  {  "device": "taxus",  "location": "1st Dg",  "segment_code": ["9"],  "diameter_mm": 2.75,  "length_mm": 32  },  {  "device": "xience",  "location": "dLCx",  "segment_code": ["13"],  "diameter_mm": 3.0,  "length_mm": 33  }  ],  "current_stents_rearrange": [  {  "device": "Onyx",  "location": "LM to pLAD",  "segment_code": ["5", "6"],  "diameter_mm": 3.5,  "length_mm": 18  }  ]  }  ``` |
| **(B) Case of which fractional flow reserve (FFR) information was well identified in JSON report.** | |
| ****Unstructured Text****  *Date of CAG PCI*:  *Coronary Angiography*  [**Findings**  Rt RA c/ 6Fr sheath  RCA c/ JR  no sig LN of RCA  LCA c/ JL  diffuse ecc. 60% LN of p-to-dLAD  no sig LN of LCx  FFR baseline 0.94 hyperemia 0.8  JL guiding -> XB guiding  sion 2EA  preballoon 2.0x15  IVUS +  Elunir 2.75x28, 3.5x33  adjuvant 4.0x15  IVUS +  post FFR baseline 0.97 hyperemia 0.94  Conclusion:**  ELSA reg [24.1.18]  UA  s/p PCI of pmLAD (Elunir 3.5x33, 2.75x28) [24.1.18] FFR 0.94/0.8 // 0.97/0.94]  *PCI *  [  Diagnosis before procedure: Coronary (artery) atherosclerosis  **Target lesion 1**{  location: mLAD  Guiding catheter: JL 6Fr-3.5 -> EBU6Fr  Guidewire: Asahi sion blue  Preballoon: Conqueror 2.0*15  Adjuvant balloon: stent balloon(10:52)  DEB:  Stent: Elunir2.75*28 (10:42)}  **Target lesion 2**{  location: pLAD  Guiding catheter: JL 6Fr-3.5 -> EBU6Fr  Guidewire: Asahi sion blue  Preballoon: Conqueror 2.0*15  Adjuvant balloon: Selethru NC 4.0*15 (10:47)  DEB:  Stent: Elunir 3.5*33 (10:44)}  **Target lesion 3**{  location:  Guiding catheter:  Guidewire:  Preballoon:  Adjuvant balloon:  DEB:  Stent:}  **Target lesion 4**{  location:  Guiding catheter:  Guidewire:  Preballoon:  Adjuvant balloon:  DEB:  Stent:}** Diagnosis after procedure:  **Coronary (artery) atherosclerosis] | ****Structured JSON****  ```json  {  "coronary_angiography": {  "catheters": {  "RCA_catheter": ["JR"],  "LCA_catheter": ["JL"]  },  "lesions": [  {  "lesion_characteristics": ["diffuse", "eccentric"],  "location": "pLAD~dLAD",  "segment_code": ["6", "7", "8"],  "luminal_narrowing_percentage": 60  }  ],  "FFR_details": [  {  "location": "pLAD~dLAD",  "segment_code": ["6", "7", "8"],  "guide_catheter": "JL",  "guide_wire": ["sion"],  "FFR_baseline": 0.94,  "drug": "adenosine",  "FFR_hyperemia": 0.8  }  ]  },  "pci_details": [  {  "target_vessel": "LAD",  "guide_catheter": ["JL 6Fr-3.5", "EBU 6Fr"],  "guide_wire": ["Asahi sion blue"],  "all_balloons": [  {  "device": "Conqueror",  "location": "mLAD",  "segment_code": ["7"],  "diameter_mm": 2.0,  "length_mm": 15  },  {  "device": "Conqueror",  "location": "pLAD",  "segment_code": ["6"],  "diameter_mm": 2.0,  "length_mm": 15  },  {  "device": "Selethru NC",  "location": "pLAD",  "segment_code": ["6"],  "diameter_mm": 4.0,  "length_mm": 15  }  ],  "stents": [  {  "device": "Elunir",  "location": "mLAD",  "segment_code": ["7"],  "diameter_mm": 2.75,  "length_mm": 28,  "pressure_atm": 10.42  },  {  "device": "Elunir",  "location": "pLAD",  "segment_code": ["6"],  "diameter_mm": 3.5,  "length_mm": 33,  "pressure_atm": 10.44  }  ],  "IVUS_used": true  }  ],  "FFR_details": [  {  "location": "pLAD~dLAD",  "segment_code": ["6", "7", "8"],  "guide_catheter": "JL",  "guide_wire": ["sion"],  "FFR_baseline": 0.94,  "drug": "adenosine",  "FFR_hyperemia": 0.8  },  {  "location": "pLAD~dLAD",  "segment_code": ["6", "7", "8"],  "guide_catheter": "JL",  "guide_wire": ["sion"],  "FFR_baseline": 0.97,  "drug": "adenosine",  "FFR_hyperemia": 0.94  }  ],  "previous_stents_rearrange": [],  "current_stents_rearrange": [  {  "device": "Elunir",  "location": "mLAD",  "segment_code": ["7"],  "diameter_mm": 2.75,  "length_mm": 28  },  {  "device": "Elunir",  "location": "pLAD",  "segment_code": ["6"],  "diameter_mm": 3.5,  "length_mm": 33  }  ]  }  ``` |

### **Supplementary Table 1. Accuracy Comparison for the Analysis on 12 Key Points by the Framework and Two Cardiologists in Internal Test.**

The total number of CAG reports including CAG information or PCI information, and the number of N/A cases, TRUE cases, and accurate samples are presented to aid in interpreting the accuracy percentages. CTO: chronic total occlusion; Cardio 1: Cardiologist 1; Cardio 2: Cardiologist 2.

| **Key Point** | **Number of N/A cases** | **Number of TRUE cases** | **Number of Accurate Samples** | | | **p-value (Framework vs Cardio 1)** | **p-value (Framework vs Cardio 2)** |
| --- | --- | --- | --- | --- | --- | --- | --- |
|  |  |  | **Framework** | **Cardio 1** | **Cardio 2** |  |  |
| **CAG Information (N = 96)** | | | | | | | |
| Location of Previous Stents | **69** | - | **96** | **88** | **87** | **0.007** | **0.003** |
| Previous Stents Details | 69 | - | 96 | 92 | 91 | 0.121 | 0.059 |
| Location and Type of Lesion | 0 | - | **94** | **78** | **77** | **<0.001** | **<0.001** |
| Anatomical Diagnosis | 0 | - | 96 | 96 | 96 | 1 | 1 |
| **PCI Information (N = 44)** | | | | | | | |
| Multivessel PCI | - | 6 | 44 | 44 | 44 | 1 | 1 |
| ≥3 Lesions Treated | - | 11 | 43 | 43 | 43 | 1 | 1 |
| Bifurcation with ≥2 Stents | - | 2 | 44 | 43 | 43 | 1 | 1 |
| ≥3 Stents Implanted | - | 5 | 43 | 43 | 43 | 1 | 1 |
| CTO PCI | - | 10 | 44 | 44 | 44 | 1 | 1 |
| Total Stent Length >60 mm | - | 2 | 43 | 43 | 43 | 1 | 1 |
| Complex PCI | - | 20 | 43 | 43 | 44 | 1 | 1 |
| Current Stents Details | 4 | - | 42 | 40 | 39 | 0.676 | 0.434 |

### **Supplementary Table 2. Qualitative Analysis of Errors (A) the Framework Made and (B) Two Cardiologists Made in Internal Test Set.**

(A) Framework Error

| **Key Point** | **Snippet of Report** | **Framework’s**  **Answer** | **Accurate**  **Answer** | **Reason for Error** |
| --- | --- | --- | --- | --- |
| **CAG-related Key Information Points** | | | | |
| Location of Lesion | Diffuse ecc. 70% LN of m-dLAD & 2nd Dg os. c severe calcification | [['3','C'], ['4','B2'], ['6','B2'], ['7','C'], ['10','C'], ['12a','A']] | [['3','C'], ['4','B2'], ['6','B2'], ['7','C'], ['8','C'], ['10','C'], ['12a','A']] | Missed dLAD lesion ‘8’ |
| Type of Lesion | Discrete eccentric 50% LN of LAD ostium | [['6','B1']] | [['6','B2']] | Identified the lesion of ‘6’ as type B1, which should be type B2 because of 'eccentric and 'ostium'. |
| **PCI-related Key Information Points** | | | | |
| Three or More Stents Implanted | Stent :  Biofreedom 2.5*14 → lesion pass failed  → Biomatrix 2.5*14 → lesion pass failed  Double 014" G/W : Dg+ Dg  Stent :  Biofreedom 2.5*14 → lesion pass failed  → Biomatrix 2.5*14 → lesion pass failed | TRUE | FALSE | The stent couldn't pass the lesion despite four attempts, which implies the stent implantation failure. But LLM counted this as four stents implanted. |
| CTO_PCI | RCA c JR 5-4  Total occlusion of pRCA (TIMI 0)  (…)  [PCI at RCA] | FALSE | TRUE | The PCI section did not mention 'CTO PCI' explicitly, but the CAG section mentioned about total occlusion lesion and PCI was done on the lesion, which requires thorough inspection of entire report. This led to judging complex PCI as FALSE. |
| Complex PCI |  | FALSE | TRUE |  |
| Current Stent Information | Stent : Xience Alpine 2.75*18 (10 atm) at LIMA-dLAD anastomosis site] | [['XienceAlpine', 2.75, 18, ['6']]] | [['XienceAlpine', 2.75, 18, ['8']]] | This patient had coronary artery bypass graft surgery, which makes the 'segment code' of the lesions complicated. LLM failed to capture the LIMA-dLAD lesion into segment code, which was inevitable. |
|  | Stent : Promus Element 3.0 *38 (16atm) at LM -mLAD | [['PromusElement', 3.0, 38, ['5','7']]] | [['PromusElement', 3.0, 38, ['5','6','7']]] | The current stent was implanted in the 'LM~mLAD' region, but LLM only identified LM and mLAD, leaving out the segment code for pLAD. Although pLAD was not explicitly mentioned, it is implicitly obvious for cardiologists. |

(B) Cardiologist Error

| **Key Point** | **Snippet of Report** | **Cardiologist’s**  **Answer** | **Accurate**  **Answer** | **Reason for Error** |
| --- | --- | --- | --- | --- |
| **CAG-related Key Points** | | | | |
| Location of Previous Stent | Patent previous stent at 3rd OM (Coronary stent 2.5*14) | [‘14’] | ['12b'] | Labeled the segment code of 3rd OM (12b) as left PL(14). |
|  | 30% NIH at pRCA (USA, unknown stent) | ['2','3','16'] | ['1','3','16','11','13'] | Labeled the segment code of pRCA (1) as mRCA (2). |
|  | 70% ISR at LM~pLAD (Xience prime 3.5*28, 3.0*33 → ISR → DEB) | ['5','6','7'] | ['5','6'] | Identified segment code of LM ~ pLAD (5, 6) as LM ~ mLAD (5,6,7) |
|  | Patent previous stent at far dLCx (other hospital) | ['13'] | ['15'] | Labeled far dLCx(15) as dLCx(13). |
|  | Total occlusion of pLCx (2003 s/p PTCA c stent at dLCx)  Total occlusion of pLAD (2003 s/p PTCA c stent at mLAD) | ['3','6','11'] | ['3','13','7'] | Labeled segment code of dLCx (13) and mLAD (7) as pLCx (11) and pLAD (6) |
|  | Patent previous stent at m-dRCA(Xience xpedition 3.5*33) | ['7','1','2','3'] | ['7','2','3'] | Absent segment code (1) was added. |
|  | 50% ISR at far dLCx (Cypher 2.5*23) | ['7','13'] | ['7','15'] | Labeled far dLCx(15) as dLCx(13). |
|  | 50% ISR at early OM (Mac 3.5*10) | ['11','6','7'] | ['6','7','11','12a'] | Omitted the segment code 12a. |
| Location of Lesion | Tubular ecc. 30% LN of 2nd OM os. | [['12','C'], ['8','C']] | [['12a','C'], ['8','C']] | Labeled the segment code of OM (12a) as RI(12). |
|  | diffuse ecc. 70% LN of d~far dLCx, small caliber | [['13','C']] | [['13','C'], ['15','C']] | Labeled only the lesion of dLCx(13), missing 'far dLCx' (15). |
|  | RCA c JR 5-4  tubular eccentric 90% LN of dRCA & PD bifurcation | [['2','B2'], ['3','B2'], ['15','B2'], ['6','B2']] | [['2','B2'], ['3','B2'], ['4','B2'], ['6','C']] | Labeled PD from RCA (4) as PD from LCA (15). |
|  | DIffuse irr 80% LN of LCx proper, OM | [['12a','C']] | [['11','C'], ['12a','C']] | Missed segment code '11'. |
|  | Discrete eccentric 50% LN of LAD ostium | [['5','B2']] | [['6','B2']] | Labeled LAD os (6) as LM(5). |
|  | Diffuse ecc. 80% LN of pLCx | ['1','C'], ['3','A'], ['16','C'], ['7','B2'] | [['1','C'], ['3','B1'], ['16','C'], ['11','C'], ['7','B2']] | Omitted segment code '11'. |
|  | 70% ISR at LM~pLAD (Xience prime 3.5*28, 3.0*33 → ISR → DEB) | [['1','B2'], ['4','C'], ['5','B2'], ['6','B2'], ['7','B2'], ['11','B2']] | [['1','B2'], ['4','C'], ['5','B2'], ['6','B2'], ['11','B2']] | Identified segment code of LM ~ pLAD (5, 6) as LM ~ mLAD (5,6,7) |
|  | 50% ISR at far dLCx (Cypher 2.5*23) | [['2','B2'], ['4','B1'], ['6','C'], ['9','B2'], ['13','B2']] | [['2','B2'], ['4','B2'], ['6','C'], ['9','B2'], ['15','B2']] | Labeled far dLCx(15) as dLCx(13). |
| Type of Lesion | discrete ecc. 80% LN of early OM os | [['12a','B1'], ['6','B2'], ['8','B2'], ['10','C'], ['1','B2'], ['2','B2']] | [['12a','B2'], ['6','B2'], ['8','B2'], ['10','C'], ['1','B2'], ['2','B2']] | Labeled the lesion ‘12a’ as type B1, identifying 'eccentric' but missing the 'os' as type B characteristic. |
|  | Total occlusion of early Dg. (TIMI 1) | [['9','B2']] | [['9','C']] | Identified lesion ‘9’ as type B2, which should be type C. |
|  | Diffuse ecc. 70% LN of OM | [['1','C'], ['9','B2'], ['7','B2'], ['8','C'], ['12a','B2']] | [['1','C'], ['9','B2'], ['7','B2'], ['8','C'], ['12a','C']] | Identified lesion ‘12a’ as type B2, which should be type C. |
|  | Discrete ecc. 80% LN of DG os. | [['1','B2'], ['9','B1'], ['11','B2']] | [['1','B2'], ['9','B2'], ['11','B2']] | Identified lesion ‘9’ as type B1, which should be type B2 because of 'eccentric' and 'os'. |
|  | Discrete ecc. 80% LN of dRCA | ['1','C'], ['3','A'], ['16','C'], ['7','B2'] | [['1','C'], ['3','B1'], ['16','C'], ['11','C'], ['7','B2']] | Identified lesion ‘3’ as type A lesion, which should be type B1 because of 'eccentric'. |
|  | Discrete ecc. 90% LN of 1st Dg os c hazziness | [['9','B1'],['11','B1']] | [['9','B2'],['11','B1']] | Identified lesion ‘’ as type B1, which should be type B2 because of 'eccentric' and 'os'. |
|  | Discrete ecc. 90% LN of mRCA | [['2','A'], ['3','C'], ['6','C'], ['7','C']] | [['2','B1'], ['3','C'], ['6','C'], ['7','C']] | Identified lesion ‘2’ as type A lesion, which should be type B1 becasuse of 'eccentric'. |
|  | Tubular ecc. 50% LN of PD os | [['2','B2'], ['4','B1'], ['6','C'], ['9','B2'], ['13','B2']] | [['2','B2'], ['4','B2'], ['6','C'], ['9','B2'], ['15','B2']] | Identified lesion ‘4’ as type B1 lesion, which should be type B2 because of 'eccentric' and 'os'. |
|  | Discrete ecc. 80% LN of 1st Dg os | [['9','B1'], ['11','B2'], ['12b','B2'], ['13','B2'], ['1','B2'], ['2','C'], ['3','C'], ['4','C']] | [['9','B2'], ['11','B2'], ['12b','B2'], ['13','B2'], ['1','B2'], ['2','C'], ['3','C'], ['4','B2']] | Identified lesion ‘9’ as type B1, which should be type B2 because of 'eccentric' and 'os'. |
|  | Discrete ecc. 50% LN of PD os | [['9','B1'], ['11','B2'], ['12b','B2'], ['13','B2'], ['1','B2'], ['2','C'], ['3','C'], ['4','C']] | [['9','B2'], ['11','B2'], ['12b','B2'], ['13','B2'], ['1','B2'], ['2','C'], ['3','C'], ['4','B2']] | Identified lesion ‘4’ ' as type C, which should be type B2." |
|  | Total occlusion of mRCA (TIMI 1) | [['2','B2'], ['3','C'], ['11','B2'], ['12a','C'], ['13','C'], ['6','C'], ['7','C']] | [['2','C'], ['3','C'], ['11','B2'], ['12a','C'], ['13','C'], ['6','C'], ['7','C']] | Identified lesion ‘2’ as type B2, which should be type C because of 'total occlusion'. |
|  | Discrete ecc 90% LN of pRCA | [['1','A'], ['2','B2'], ['12a','B2']] | [['1','B1'], ['2','B2'], ['12a','B2']] | Identified lesion ‘1’ as type A, which should be type B1 because of 'eccentric'. |
|  | 70% ISR at p-mLAD (Radius 4.0*20) c severe calcification | [['3','C'], ['4','B2'], ['6','C'], ['7','C'], ['8','C'], ['10','C'], ['12a','A'] | [['3','C'], ['4','B2'], ['6','B2'], ['7','C'], ['8','C'], ['10','C'], ['12a','A']] | Identified ISR lesion ‘6’ as type C, which should be type B2" |
|  | Discrete ecc. 50% LN of PD os. | [['3','B2'], ['4','A']] | [['3','B2'], ['4','B1']] | Identified lesion ‘4’ as type B1, which should be type B2 because of 'eccentric' and 'os'. |
| **PCI-related Key Points** | | | | |
| Three or More Lesions Treated | [PCI at RCA]  SAL 6-1 SH  014" G/W: Pilot  Balloon: PCI balloon 2.5*15  Stent: Biofreedom 3.0*33 (6atm) at p~mRCA  f/u angio : 30%< R/S  Adjuvant balloon : NC 3.0*12 (20atm)  f/u angio : No R/S  [PCI at LAD]  XBG 6-3.5  014" G/W  Balloon: PCI balloon 2.5*15  Stent: Biofreedom 3.0*28 (8atm) at dLAD  Biofreedom 3.5*33 (8atm) at mLAD  Adjuvant balloon : NC 3.0*12 (24atm)  f/u angio : No R/S | FALSE | TRUE | There were two PCI sections (RCA, LAD), but cardiologist saw only the latter one, which led to identifying the two key criteria (‘three or more lesions treated’ and ‘three or more stents implanted’) above as FALSE. It propagated to identifying complex PCI as FALSE. |
| Three or More Stents Implanted |  | FALSE | TRUE |  |
| Complex PCI |  | FALSE | TRUE |  |
| CTO_PCI | RCA c JR 5-4  Chronic total occlusion of pRCA(TIMI 0)  (…)  [PCI at RCA] | FALSE | TRUE | Missed CTO PCI |
| Bifurcation with Two or More Stents | Stent : Resolute integrity 2.5*26 (12atm) at p~dLCx  (…)  Stent : Resolute integrity 3.0*30 (9atm) at LM~pLAD | FALSE | TRUE | Missing p-LCx segment code of first stent led to labeling as FALSE. |
| Current Stent Information | Stent : Orsiro 3.5*18 at LCX os~pLCX (TAP technique) | ['Orsiro', '3.5', '18', ['5','11']] | [['Orsiro', 3.5, 18, ['11']]] | Identified the segment code of current stent as '5' and '11', which should be only '11'. |
|  | [PCI: LCX proper]  (…)  Stent: Genoss 2.5*28→ R/S>20%, underexpansion | [['Genoss'2.5, 28,['12a']]] | [['Genoss',2.5, 28,['11']]] | Identified segment code of current stent as OM (12a), which should be pLCx (11). |
|  | Stent :  Biofreedom 2.5*14 → lesion pass failed  → Biomatrix 2.5*14 → lesion pass failed  Double 014" G/W : Dg+ Dg  Stent :  Biofreedom 2.5*14 → lesion pass failed  → Biomatrix 2.5*14 → lesion pass failed | [['Biofreedom', 2.5,14,['9']]] | N/A | The stent couldn't pass the lesion but identified as implanted stent. |

### **Supplementary Table 3. Accuracy Comparison for the Analysis on 12 Key Points by the Framework and Two Cardiologists in External Test.**

The total number of CAG reports including CAG information or PCI information, and the number of N/A cases, TRUE cases, and accurate samples are presented to aid in interpreting the accuracy percentages. CTO: chronic total occlusion.

| **Key Point** | **Number of N/A cases** | **Number of TRUE cases** | **Number of Accurate Samples of the Framework** |
| --- | --- | --- | --- |
| **CAG Information (N = 98)** | | | |
| Location of Previous Stents | 81 | - | 94 |
| Previous Stents Details | 81 | - | 96 |
| Location and Type of Lesion | 0 | - | 93 |
| Anatomical Diagnosis | 0 | - | 94 |
| **PCI Information (N = 58)** | | | |
| Multivessel PCI | - | 2 | 58 |
| ≥3 Lesions Treated | - | 9 | 58 |
| Bifurcation with ≥2 Stents | - | 3 | 57 |
| ≥3 Stents Implanted | - | 4 | 58 |
| CTO PCI | - | 8 | 58 |
| Total Stent Length >60 mm | - | 2 | 58 |
| Complex PCI | - | 20 | 58 |
| Current Stents Details | 8 | - | 56 |

### **Supplementary Table 4. Qualitative Analysis of Errors the Framework Made in External Test Set.**

| **Key Point** | **Snippet of Report** | **Framework’s**  **Answer** | **Accurate**  **Answer** | **Reason for Error** |
| --- | --- | --- | --- | --- |
| **CAG-related Key Points** | | | | |
| Location of Previous Stent | s/p PTCA with Ultimaster 2.75x28 at LAD ISR (type IB) | ['7'] | N/A | The current stent has likely been recorded as the previous stent because of the 's/p (status post)' notation preceding it. |
|  | s/p PCI of pRCA (ultimaster 3.5x33) [24.3.20]  s/p PCI of dRCA (ultimaster 2.75x24), OM (Elunir 2.75x28) [24.3.22]] | ['1', '3', '12b'] | N/A | The current stent has likely been recorded as the previous stent because of the 's/p (status post)' notation preceding it. |
|  | Patent stent at pLAD | ['1'] | ['1', ‘6’] | Patent stent in pLAD was overlooked. |
|  | s/p PTCA with Cre8 3.5x 20 at pLAD (stent protrusion LM) | [‘6’] | N/A | The current stent has likely been recorded as the previous stent because of the 's/p (status post)' notation preceding it. |
| Previous Stent Information | s/p PTCA with Ultimaster 2.75x28 at LAD ISR (type IB) | [['Ultimaster', 2.75, 28, ['7']]] | N/A | The current stent has likely been recorded as the previous stent because of the 's/p (status post)' notation preceding it. |
|  | s/p POBA with DEB(sequent) 3.0x30 at mLAD ISR | [['DEB (Sequent)', 3.0, 30, ['7']]] | N/A | "The currently treated DCB has been mistakenly interpreted as the previously treated stent." |
| Location of Lesion | near total occlusion of PL br. c TIMI 2 distal flow | [['7', 'C'], ['11', 'B2']] | [['7', 'C'], ['11', 'B2'], ['16','B2']] | The information about the PL from RCA (16 segment) was missed. |
|  | 90% LN of LCx  --> PCI with stent at LCx  (…)  **Target lesion 1**{  location: dLCx  Guiding catheter: JL6Fr-3.5->EBU6Fr  Guidewire: BMW  Preballoon: Genoss 2.0*15  Adjuvant balloon:  DEB:  Stent: Genoss 2.75*33 (12:13)} | [['4', 'A']] | [['4', 'A'], ['13','A']] | In the CAG section, it only states '90% LN LCx', so it is unclear whether it refers to dLCx. However, it had to be inferred from the PCI section where a stent was placed in the dLCx. |
| Type of Lesion | diffuse heavy calcification of RCA | [['1', 'C'], ['2', 'C'], ['3', 'C'], ['4', 'C']] | [['3', 'B2'], ['4', 'C']] | 'diffuse heavy calcification' was interpreted as 'diffuse' lumen narrowing and 'heavy calcification'. This led to marking the entire RCA segments as diffuse lumen narrowing lesions. |
|  | revealed 70% ISR of prev. LAD stent | [['1', 'A'], ['5', 'C'], ['11', 'C'], ['7', 'B2']] | [['1', 'A'], ['5', 'C'], ['11', 'C'], ['6','B2'], ['7', 'B2']] | ISR is also present in the pLAD, but it was overlooked. |
|  | ISR prev. pRCA stent (taxus 3.5x20) [06.4.17] | [['6', 'B2']] | [['6', 'B2'], ['1','B2']] | ISR is present in the pRCA, but it was overlooked. |
| Anatomical Diagnosis | near total occlusion of PL br. c TIMI 2 distal flow | 2VD | 3VD | The information about the PL from the RCA (16 segment) is missing in the CAG, which led to missing the presence of a lesion in the right coronary artery. As a result, it was assessed as 2VD instead of 3VD. |
|  | Patent stent at pLAD | 1VD | 2VD | Patent stent in pLAD was overlooked. As a result, it was assessed as 1VD instead of 2VD. |
|  | no sig coronary stenosis  [provocadtion test]  RCA ->E2 + (70% stenosis at pRCA ,ST change + ) | 1VD | N/A | During the provocation test on the pRCA, vasospasm occurred, but the anatomical diagnosis was normal. As a result, it was assessed as 1VD instead of ‘N/A’. |
|  | 90% LN of LCx  --> PCI with stent at LCx  (…)  **Target lesion 1**{  location: dLCx  Guiding catheter: JL6Fr-3.5->EBU6Fr  Guidewire: BMW  Preballoon: Genoss 2.0*15  Adjuvant balloon:  DEB:  Stent: Genoss 2.75*33 (12:13)} | 1VD | 2VD | In the CAG section, it only states '90% LN LCx', so it is unclear whether it refers to dLCx. However, it had to be inferred from the PCI section where a stent was placed in the dLCx. . As a result, it was assessed as 1VD instead of 2VD. |
| **PCI-related Key Points** | | | | |
| Bifurcation with Two or More Stents | Conclusion:Successful PCI with stents at p- to m-LAD, big Dx  (…)  **Target lesion 2**{  location: mLAD  Guiding catheter: JL 6F-3.5  Guidewire: Pilot, Asahi sion black  Preballoon: GENOSS 2.5*15  Adjuvant balloon: Euphora NC 3.5*15 (14:23)  DEB:  Stent: Xience Skypoint 3.0*33 (13:58)} | FALSE | TRUE | In the earlier part where the CAG section and PCI section were mixed, it was stated that two stents were placed in the p-mLAD and big Dx. However, in the separately organized PCI section below, the placement of the stent in the pLAD was omitted, leading LLM to make its judgment based on the latter PCI section. |
| Current Stent Information | Near total occlusion of d-LCx c TIMI 2 distal flow  -> POBA 2.0x15  -> DES (Ultimaster 2.75x24) implantation  Conclusion:Successful PCI with stent at d-LCx  (…)  **Target lesion 1**{  location: pLCX  Guiding catheter: JL 6F-3.5  Guidewire: BMW  Preballoon: GENOSS 2.0*15  Adjuvant balloon:  DEB:  Stent: Ultimaster 2.75*24 (10:17)} | [['Ultimaster', 2.75, 24, ['11']]] | [['Ultimaster', 2.75, 24, ['13']]] | The CAG section and the conclusion stated that the stent was successfully implanted in the dLCx. But the PCI section listed the stent location as pLCx which was likely a typo. It seems that the error was due to focusing on the directly indicated 'location' section. |
